# A placebo-controlled double blind trial of hydroxychloroquine in mild-to-moderate COVID-19

**DOI:** 10.1101/2020.10.19.20214940

**Authors:** Vincent Dubée, Pierre-Marie Roy, Bruno Vielle, Elsa Parot-Schinkel, Odile Blanchet, Astrid Darsonval, Caroline Lefeuvre, Chadi Abbara, Sophie Boucher, Edouard Devaud, Olivier Robineau, Patrick Rispal, Thomas Guimard, Emma d’Anglejean, Sylvain Diamantis, Marc-Antoine Custaud, Isabelle Pellier, Alain Mercat, for the HYCOVID study group

**Affiliations:** Service des Maladies Infectieuses et Tropicales, CHU d’Angers, Angers, France; CRCINA, Inserm, Université de Nantes, Université d’Angers, Angers, 44200 Nantes, France; Emergency Department, CHU d’Angers, Angers, France; Institut MitoVasc, UMR CNRS 6215 INSERM 1083, Université d’Angers, Angers, France; Biostatistics and Methodology Department, Maison de la recherche, CHU d’Angers, France; Centre de Ressources Biologiques, BB-0033-00038, Angers, CHU d’Angers, France; Service Pharmacie, CHU d’Angers, Angers, France; Département des Agents Infectieux, Laboratoire de virologie, CHU Angers, Angers, France; Service d’ORL et Chirurgie Cervico-faciale, CHU d’Angers, Angers, France; Institut MitoVasc, UMR CNRS 6215 INSERM 1083, Université d’Angers, Angers, France; Service de médecine interne et maladies infectieuses, CH R. Dubos, Pontoise, France; Service Universitaire des Maladies Infectieuses et du Voyageur, CH de Tourcoing, Tourcoing, France; Service de médecine interne, CH Agen, Agen, France; Service de médecine Post-urgence, CH Départemental de Vendée, La Roche sur Yon, France; Service de médecine interne et maladies infectieuses, CH Versailles - Hôpital André Mignot, Le Chesnay, France; Service de médecine polyvalente et maladies infectieuses, Groupe Hospitalier Sud Ile de France, Melun, France; Centre de Recherche Clinique, CHU d’Angers, Angers, France; MITOVASC Institute, UMR CNRS 6015, UMR INSERM 1083, Angers, France; Unité d’hématologie et d’oncologie pédiatrique, CHU d’Angers, Inserm U1232-CRCINA, Université d’Angers, Angers, France; Département de Médecine Intensive-Réanimation, CHU d’Angers, Université d’Angers, Angers, France

**Keywords:** COVID-19, hydroxychloroquine, randomized controlled trial, SARS-CoV-2

## Abstract

**Background:** The efficacy of hydroxychloroquine in coronavirus disease 2019 (COVID-19) remains controversial.

**Methods:** We conducted a multicentre randomized double-blind placebo-controlled trial evaluating hydroxychloroquine in COVID-19 patients with at least one of the following risk factors for worsening: age ≥75 years, age between 60 and 74 years, and presence of at least one comorbidity, or need for supplemental oxygen (≤3 L/min). Eligible patients were randomized in a 1:1 ratio to receive either 800mg hydroxychloroquine on Day 0 followed by 400mg per day for 8 days or a placebo. The primary endpoint was a composite of death or tracheal intubation within 14 days following randomization. Secondary endpoints included mortality and clinical evolution at Day 14 and 28, viral shedding at Day 5 and 10.

**Results:** The trial was stopped after 250 patients were included due to a slowdown of the pandemic in France. The intention-to-treat population comprised 123 and 124 patients in the placebo and hydroxychloroquine groups, respectively. The median age was 77 years and 151 patients required oxygen therapy. The primary endpoint occurred in nine patients in the hydroxychloroquine group and eight patients in the placebo group (relative risk 1.12; 95% confidence interval 0.45– 2.80; P=0.82). No difference was observed between the two groups in any of the secondary endpoints.

**Conclusion:** In this trial involving mainly older patients with mild-to-moderate COVID-19, patients treated with hydroxychloroquine did not experience better clinical or virological outcomes than those receiving the placebo.

## Introduction

Severe acute respiratory syndrome coronavirus-2 (SARS-CoV-2) disease (COVID-19) has affected more than 30 million individuals and caused more than one million deaths within nine months; severe forms are more frequent in older patients and those with comorbidities, such as hypertension, cardiovascular diseases, obesity, and diabetes [1].

COVID-19 mainly affects the respiratory tract, causing diffuse alveolar damage, but also triggers coagulopathy and vascular lesions [2]. A delayed clinical worsening is frequently observed [3]. This phenomenon is assumed to be linked to an excessive immune reaction that may be attenuated by immunomodulatory agents such as corticosteroids [4-6].

Hydroxychloroquine, a derivative of chloroquine that is commonly used in certain autoimmune diseases, has been previously shown to inhibit SARS-CoV-2 replication in vitro [7].

Hydroxychloroquine has immunomodulatory, anti-inflammatory, and anti-thrombotic properties [8], which could also theoretically prevent or limit secondary inflammatory damage. A preliminary study suggested that this drug could play a role in shortening viral shedding [9]. However, clinical trials evaluating the efficacy of hydroxychloroquine in COVID-19 subsequently reported controversial and often disappointing results [10–18].

The aim of HYCOVID trial was to evaluate the efficacy and safety of hydroxychloroquine in adult patients with mild-to-moderate COVID-19 at risk of worsening.

## Patients and Methods

### Trial design

HYCOVID was a double-blind, placebo-controlled, randomized trial conducted from April 2 to May 21, 2020 in 48 hospitals in France and the Principality of Monaco. Eligible patients were randomly assigned in a 1:1 ratio to receive either hydroxychloroquine (200 mg tablets, orally) or its matching placebo at a dose of two tablets twice daily on the first day followed by one tablet twice daily for 8 days (total hydroxychloroquine dose of 4 g) plus standard care as needed.

Patients were randomized immediately after their inclusion into the study and stratified according to the following criteria: age ≥75 years, oxygen dependence, diagnostic criteria for COVID-19, initial symptoms of COVID-19 for less than 7 days, hospitalization, concomitant treatment with azithromycin, concomitant treatment with lopinavir/ritonavir, treatment with corticosteroids, and centre.

### Patients

Men and non-pregnant women aged ≥18 years with a diagnosis of COVID-19 confirmed by positive SARS-CoV-2 reverse transcriptase-polymerase chain reaction (RT-PCR) on a nasopharyngeal swab within 2 days were assessed for eligibility. In centres facing a shortage of swabs or RT-PCR material, the diagnosis could be made based on a chest computed tomography (CT) scan showing typical features of COVID-19. Patients were eligible if they had at least one of the following risk factors for worsening: (i) age ≥75 years; (ii) age between 60 and 74 years and presence of at least one of the following comorbidities: obesity (body mass index ≥30 kg/m^2^), arterial hypertension requiring treatment, or diabetes mellitus requiring treatment; (iii) need for supplemental oxygen to reach a peripheral capillary oxygen saturation of more than 94% (SpO2 >94%) or a ratio of oxygen partial pressure to fractional inspired oxygen less than or equal to 300 mmHg (PaO2/FiO2 ≤300 mmHg).

Patients requiring more than 3 L/min of oxygen to reach an SpO2 of 94% were excluded, as were those with a clinical condition necessitating admission to intensive care unit, a negative SARS-CoV-2 RT-PCR, a short-term life-threatening comorbidity (life expectancy <3 months), any condition contraindicating hydroxychloroquine treatment (known hypersensitivity or allergy, retinopathy, concomitant treatment associated with a risk of ventricular arrhythmias, use of medications that are contraindicated with hydroxychloroquine and cannot be replaced or stopped during the trial), or conditions associated with an increased risk of adverse event (see Supplementary Appendix for details).

### Clinical and laboratory monitoring

All data related to this study were collected using a secured standardized electronic case report form based on the patients’ medical records. For patients with an initial positive SARS-CoV-2 RT-PCR, nasopharyngeal swabs were sampled on Day 5 and 10 after randomization, and SARS-CoV-2 RT-PCR was performed using the same technique as the initial RT-PCR.

All adverse events occurring during a patient’s participation were declared to the sponsor, with the exception of adverse events linked to the COVID-19 infection itself. The following serious adverse events were specifically assessed: cardiac rhythm or conduction disorders, seizures, hypoglycemia, vision disorders, vomiting, rash, and pruritus.

### Outcome measures

The primary endpoint was a composite of death and the need for invasive mechanical ventilation within 14 days following randomization. Secondary efficacy outcomes included the rate of mortality or invasive ventilation within 28 days following treatment initiation, clinical improvement using the World Health Organization nine-point Ordinal Scale for Clinical Improvement for COVID-19 (with scores ranging from 0 [patients at home without any clinical or biological sign of infection] to 8 [death] [19]) at Day 14 and 28, all-cause mortality at Day 14 and 28, the rate of RT-PCR tests positive for SARS-CoV-2 on nasopharyngeal swab samples at Day 5 and Day 10, and the rate of symptomatic venous thrombo-embolic events at Day 28. Three criteria were used to assess clinical evolution: the absence of deterioration (stability or decrease of at least one point on the ordinal scale), clinical improvement (decrease of at least one point on the ordinal scale), and recovery (score of 0, 1, or 2 on the ordinal scale). The secondary safety outcome consisted in the rate of serious adverse events at Day 28. Clinical events were adjudicated by an independent event adjudication committee, whose members were unaware of group assignments.

### Trial oversight

The protocol was written by members of the steering committee (see Supplementary appendix for details). The principal investigator and the chair were responsible for all aspects related to the conduct of the study. The protocol was approved by an independent protection committee and by the French National Agency for Medicines and Health Products Safety (ANSM), according to French regulations. Written informed consent was obtained from each patient or from the patient’s legal representative if the patient was unable to provide consent. This trial was conducted in accordance with the principles of the Declaration of Helsinki and the Good Clinical Practice guidelines. The trial was supported by a grant from the French Ministry of Health, as well as by private and corporate donations (see Supplementary appendix for details). None of the funding organizations played any role in the trial design, data collection, analysis of results, or writing of the manuscript.

### Statistical analysis

Based on the first available epidemiological data on COVID-19 [20, 21], we estimated the rate of the primary outcome to be 20%. A headcount of 615 patients per group allows demonstrating, under a bilateral hypothesis, an absolute difference of 6% between the two groups (relative difference of 30%), with an alpha risk of 5% and a power of 80%. To allow for up to 5% non-evaluable or lost-to-follow-up patients, we set the sample size at 1,300 patients in total.

Our main analysis was performed in the intention-to-treat population. Interim analyses were performed by conducting the triangle test on every 50 patients and were submitted to the Independent Data and Safety Monitoring Board that was formed for this study. A sensitivity analysis was performed in the per-protocol population, i.e., patients with no major deviation from the study protocol. For the safety analysis, all patients who received at least one dose of hydroxychloroquine were included in the intervention group.

The rates of the primary outcome were compared between the two groups using the Chi-squared test. The relative risks for each clinical event at Day 14 and 28 and their 95% confidence interval (CI) were calculated with an adjustment taking into account the baseline status using a Mantel-Haenszel estimation.

A sequential analysis based on the triangle test was performed using R software, version 3.6.3. Stata software (version 13) was used for other analyses. Further details regarding the statistical analysis are provided in the statistical analysis plan, available in the Supplementary Appendix.

### Role of the funding sources

The study was partially funded by a grant from the French Ministry of Health through a national call for proposals for therapeutic trials on COVID-19. The trial also received an exceptional donation from the *Pays de la Loire* region and from the *Angers Loire Métropole* conurbation. None of the funding sources had any role in trial design, data collection, monitoring, management and analysis, or writing of the manuscript.

## Results

### Patients

The trial started on April 1, 2020 and was suspended on May 26, 2020 by the French regulatory authority because of reports of hydroxychloroquine toxicity in other trials. It was definitively stopped by the sponsor on June 9, 2020 because of a low inclusion rate in the context of the slowdown of the pandemic in France.

Of the 1,822 eligible patients in 34 of the centres that completed the screening survey, 202 (11%) were included in the study. In total, 250 patients were randomized, of which 125 were assigned to receive hydroxychloroquine and 125 the placebo (Figure 1 and Figure S1). Among them, one and two patients withdrew consent in the hydroxychloroquine and placebo groups, respectively. The per-protocol population comprised 226 patients: 116 in the placebo group and 110 in the hydroxychloroquine group. A summary of the major violations that led to exclusion from the per-protocol population is provided in the Supplementary appendix.

**Figure 1.**
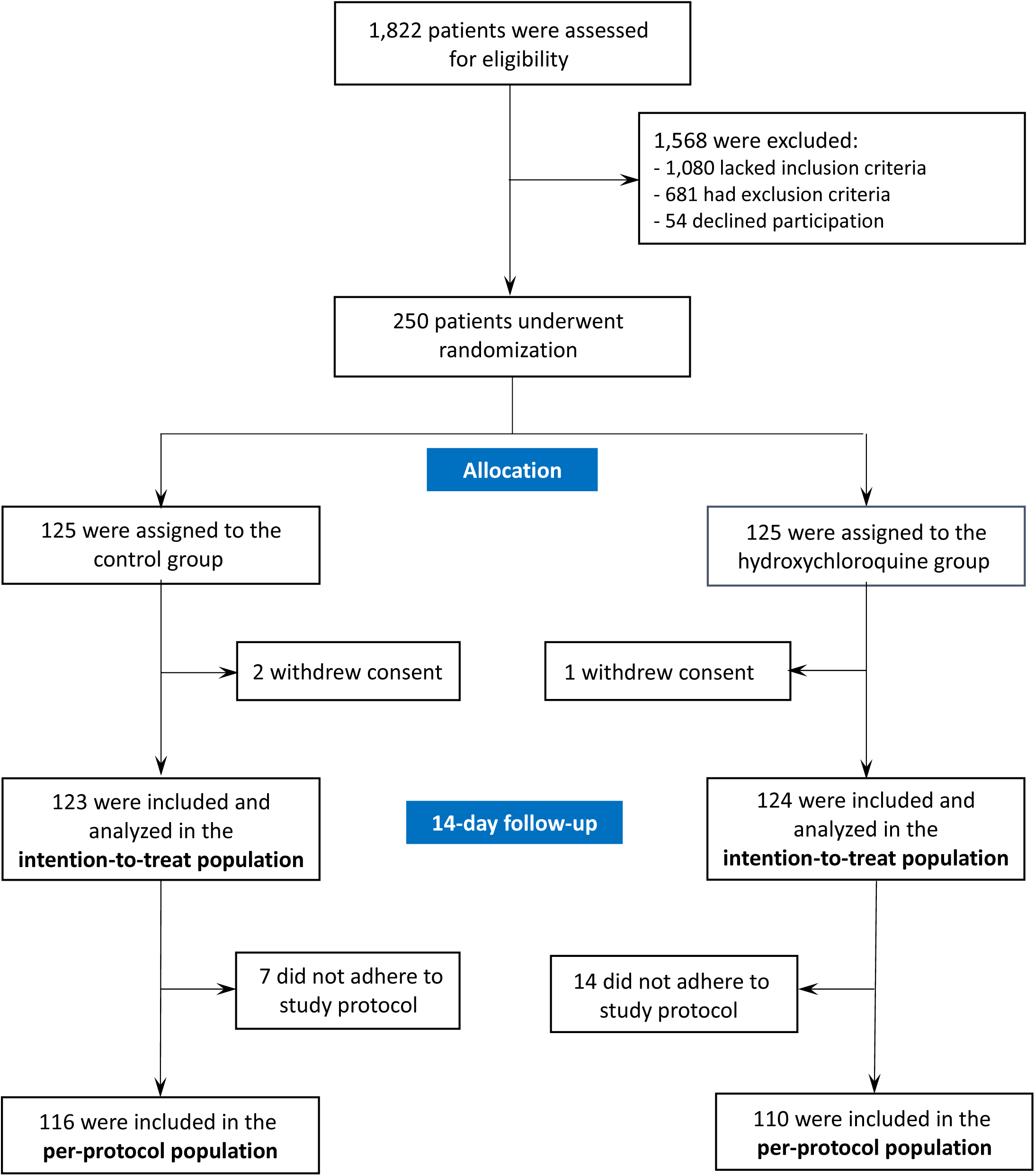
Patient selection, treatment allocation, and follow-up. Assessment of eligibility criteria was performed in 33 of the 51 participating centers, and 202 patients were included in these centers (11%).

The median age was 77 (interquartile range [IQR]: 58–86) years (Figure S2), and 60% of the patients required supplemental oxygen at baseline (Table 1). The median time from symptom onset to randomization was 5 days (IQR: 3–9). The treatment groups were well balanced for baseline demographic and clinical characteristics, as well as for the treatments of interest received at randomization.

**Table 1:** Baseline demographic, biological, and clinical characteristics of the patients

| Characteristic | All patients<br>(N = 250) | Hydroxychloroquine<br>(N = 125) | Placebo<br>(N = 125) |
| --- | --- | --- | --- |
| Median age (IQR) – yr | 77 (58-86) | 76 (60-85) | 78 (57-87) |
| Male sex – no. (%) | 121 (48.4) | 65 (52.0) | 56 (44.8) |
| Diagnostic criteria – no. (%) |  |  |  |
| Positive RT-PCR SARS-CoV-2 | 247 (98.8) | 124 (99.2) | 123 (98.4) |
| Positive CT-scan (RT-PCR not done) | 3 (1.2) | 1 (0.8) | 2 (1.6) |
| Severity risk criteria – no. (%)* |  |  |  |
| Age ≥ 75 years | 134 (53.6) | 66 (52.8) | 68 (54.4) |
| Age ≥ 60 years and comorbidity** | 1 (0.4) | 1 (0.8) | 0 |
| Supplemental oxygen requirement | 151 (60.4) | 77 (61.6) | 74 (59.2) |
| Comorbidities – no. (%) |  |  |  |
| Chronic respiratory disease | 28 (11.2) | 15 (12.0) | 13 (10.5) |
| Diabetes | 43 (17.3) | 20 (16.0) | 23 (18.6) |
| Arterial hypertension | 133 (53.4) | 65 (52.0) | 68 (54.8) |
| Heart disease | 72 (28.9) | 34 (27.2) | 38 (30.7) |
| Cerebrovascular disease | 43 (17.3) | 21 (16.8) | 22 (17.7) |
| Obesity (BMI >30 kg/m<sup>2</sup>) | 65 (27.7) | 25 (22.1) | 40 (32.8) |
| Active smoker | 6 (2.4) | 3 (2.4) | 3 (2.4) |
| Score on ordinal scale – no. (%) |  |  |  |
| 1. Ambulatory, no limitation of activity | 2 (0.8) | 1 (0.8) | 1 (0.8) |
| 2. Ambulatory, limitation of activity | 1 (0.4) | 1 (0.8) | 0 (0.0) |
| 3. Hospitalized, no oxygen therapy | 96 (34.4) | 46 (36.8) | 50 (40.0) |
| 4. Hospitalized, oxygen therapy (≤ 4 L/min) | 151 (60.4) | 77 (61.6) | 74 (59.2) |
| Concomitant treatment – no. (%) |  |  |  |
| Azithromycin | 21 (8.4) | 10 (8.0) | 11 (8.8) |
| Other antibiotics | 122 (48.8) | 62 (49.6) | 60 (48.0) |
| Lopinavir – ritonavir | 3 (1.2) | 1 (0.8) | 2 (1.6) |
| Corticosteroids | 24 (9.6) | 13 (10.4) | 11 (8.8) |
| Angiotensin-converting enzyme inhibitors or<br>angiotensin receptor blockers | 82 (33.3) | 43 (34.4) | 39 (32.2) |
| Symptoms – no. (%) |  |  |  |
| Body temperature >38°C | 53 (21.3) | 28 (22.4) | 25 (20.2) |
| Diarrhea | 34 (13.6) | 22 (17.6) | 12 (9.6) |
| Rhinitis | 16 (6.4) | 9 (7.2) | 7 (5.6) |
| Anosmia or hyposmia | 36 (14.5) | 19 (15.2) | 17 (13.7) |
| Confusion | 33 (13.2) | 16 (12.8) | 17 (13.6) |
| Median time (IQR) from onset of symptoms to inclusion<br>– days | 5 (3-9) | 5 (3-9) | 5 (3-8) |

\*One patient may have several severity risk criteria.
\*\*Arterial hypertension requiring treatment, diabetes requiring treatment, obesity with body mass index ≥30 kg/m<sup>2</sup>

### Primary outcome

There was no significant difference in the rate of the primary endpoint, which occurred within 14 days following randomization in 8 of 123 patients assigned to the placebo group (6.5%) and 9 of 124 patients assigned to the hydroxychloroquine group (7.3%) (relative risk 1.12; 95% CI: 0.45 to 2.80; P=0.82) (Table 2). At 28 days after randomization, 9.8% (12/123) of the patients in the placebo group had died or had been intubated compared to 7.3% (9/124) in the hydroxychloroquine group (relative risk 0.74; 95% CI: 0.33 to 1.70). Event-free survival analysis using the Kaplan–Meyer method did not show a significant difference between the two groups (P=0.63 by log-rank test) (Figure S3).

**Table 2:** Outcomes in the intention-to-treat population

| **Outcomes** | **All patients**<br>**(N = 247)** | **Hydroxychloroquine**<br>**(N = 124)** | **Placebo**<br>**(N = 123)** | **Relative risk (95% CI)** |
| --- | --- | --- | --- | --- |
| Primary outcome |  |  |  |  |
| Day 14 | 17 (6.9) | 9 (7.3) | 8 (6.5) | 1.12 (0.45–2.80) |
| Day 28 | 21 (8.5) | 9 (7.3) | 12 (9.8) | 0.74 (0.33–1.70) |
| Mortality |  |  |  |  |
| Day 14 | 12 (4.9) | 6 (4.8) | 6 (4.9) | 0.99 (0.33–2.99) |
| Day 28 | 17 (6.9) | 6 (4.8) | 11 (8.9) | 0.54 (0.21–1.42) |
| Use of intubation and mechanical ventilation |  |  |  |  |
| Day 14 | 6 (2.4) | 3 (2.4) | 3 (2.4) | 0.99 (0.20–4.82) |
| Day 28 | 7 (2.8) | 3 (2.4) | 4 (3.3) | 0.74 (0.17–3.26) |
| Clinical evolution, as compared to Day 0 |  |  |  |  |
| Day 14 |  |  |  |  |
| Absence of deterioration | 223 (90.3) | 112 (90.3) | 111 (90.2) | 1.01 (0.93–1.09) |
| Clinical improvement | 165 (66.8) | 84 (67.7) | 81 (65.9) | 1.01 (0.86–1.18) |
| Recovery | 139 (56.3) | 71 (57.3) | 68 (55.3) | 1.03 (0.83–1.27) |
| Day 28 |  |  |  |  |
| Absence of deterioration | 227 (91.9) | 115 (92.7) | 112 (91.1) | 1.02 (0.95–1.10) |
| Clinical improvement | 191 (77.3) | 98 (79.0) | 93 (75.6) | 1.03 (0.90–1.16) |
| Recovery | 175 (70.9) | 91 (73.4) | 84 (68.3) | 1.06 (0.91–1.24) |
| Positive SARS-CoV-2 RT-PCR<sup>1</sup> |  |  |  |  |
| At Day 5 | 148/203 (72.9) | 75/103 (72.8) | 73/100 (73.0) | 1.00 (0.84–1.18) |
| At Day 10 | 99/174 (56.9) | 52/91 (57.1) | 47/83 (56.6) | 1.01 (0.78–1.31) |
| Venous thrombo-embolic events |  |  |  |  |
| Day 28 | 4 (1.6) | 2 (1.6) | 2 (1.6) | 0.99 (0.14–6.93) |

Data are no. (%) of all patients, unless otherwise indicated.
<sup>1</sup>RT-PCR could not be performed in all patients. Data are no. of positive RT-PCR/no. of RT-PCR performed (% of positive RT-PCR)

In the per-protocol population, the primary outcome was observed in 7 of the 110 patients treated with hydroxychloroquine (6.4%) and in 6 of the 116 patients (5.2%) who received the placebo. The relative risk for death or invasive mechanical ventilation was 1.23 (95% CI: 0.43–3.55) (Table S2). No significant difference between the treatment groups was observed at Day 28.

There was no significant difference in the rate of the primary endpoint between patients assigned to placebo and those assigned to hydroxychloroquine in any of the analyzed subgroups (Figure 2).

**Figure 2.**
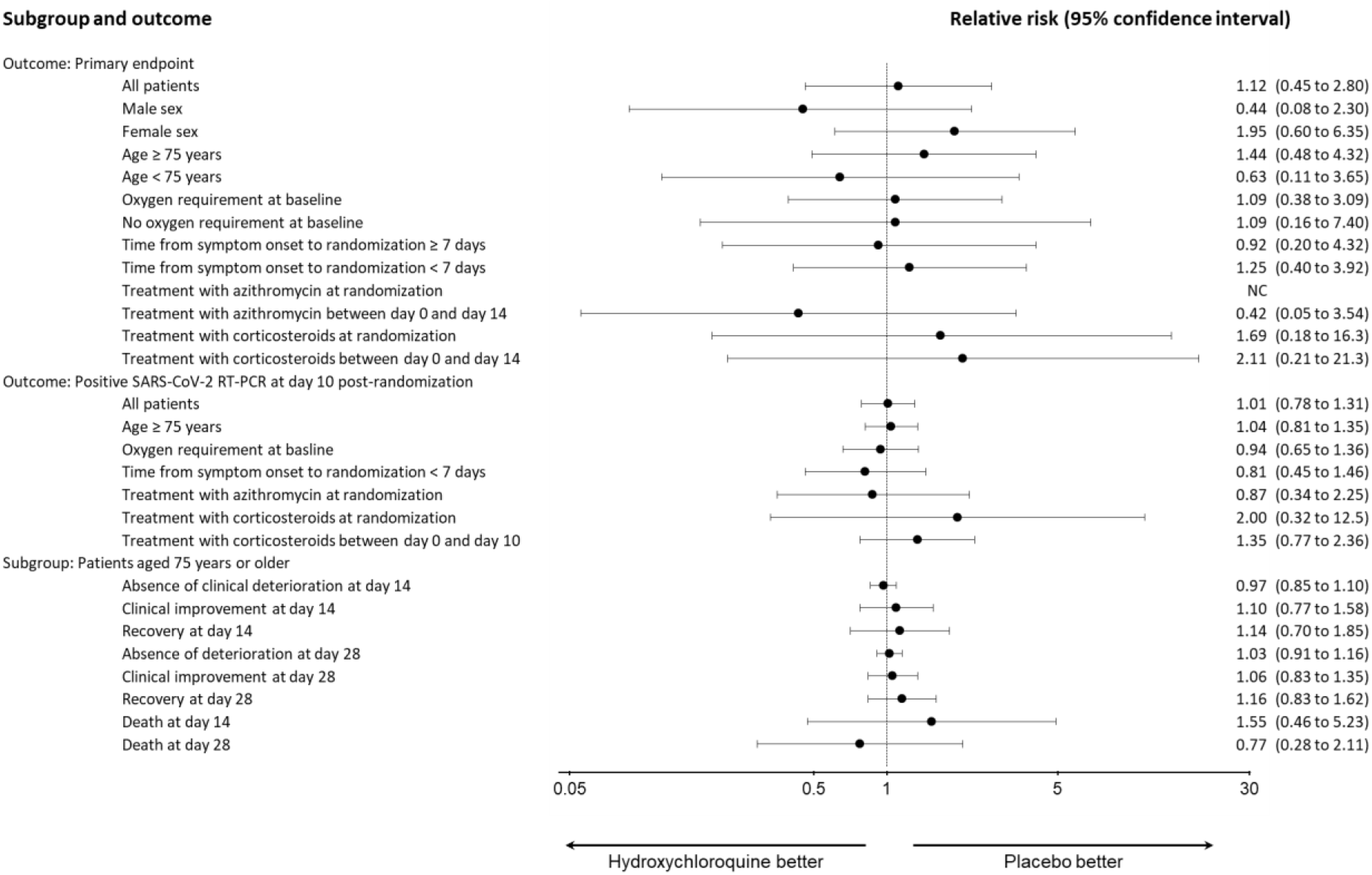
Analysis of outcomes in predefined subgroups. For analysis of the primary outcome in the subgroup of patients receiving azithromycin at randomization, the relative risk could not be calculated because the primary endpoint occurred in 0 of 10 patients who received both azithromycin and hydroxychloroquine compared to 3 of 11 patients who received azithromycin and the placebo.

### Secondary outcomes

The clinical evolution assessed by the change in ordinal scale score between Day 0 and Day 14 and between Day 0 and 28 did not differ between the two groups (Table 2 and Figure 3). At Day 14, 68 of the 123 patients assigned to the placebo had returned home (55.3%) compared to 71 of the 124 patients assigned to hydroxychloroquine (57.3%). The rate of clinical improvement was 65.9% (81/123) in the placebo group and 67.7% (84/124) in the hydroxychloroquine group. At Day 28, the rate of clinical improvement was 75.6% (93/123) and 79.0% (98/124) in the placebo and hydroxychloroquine groups, and 68.3% (84/123) and 73.4% (91/124) of patients had been discharged from hospital, respectively.

**Figure 3.**
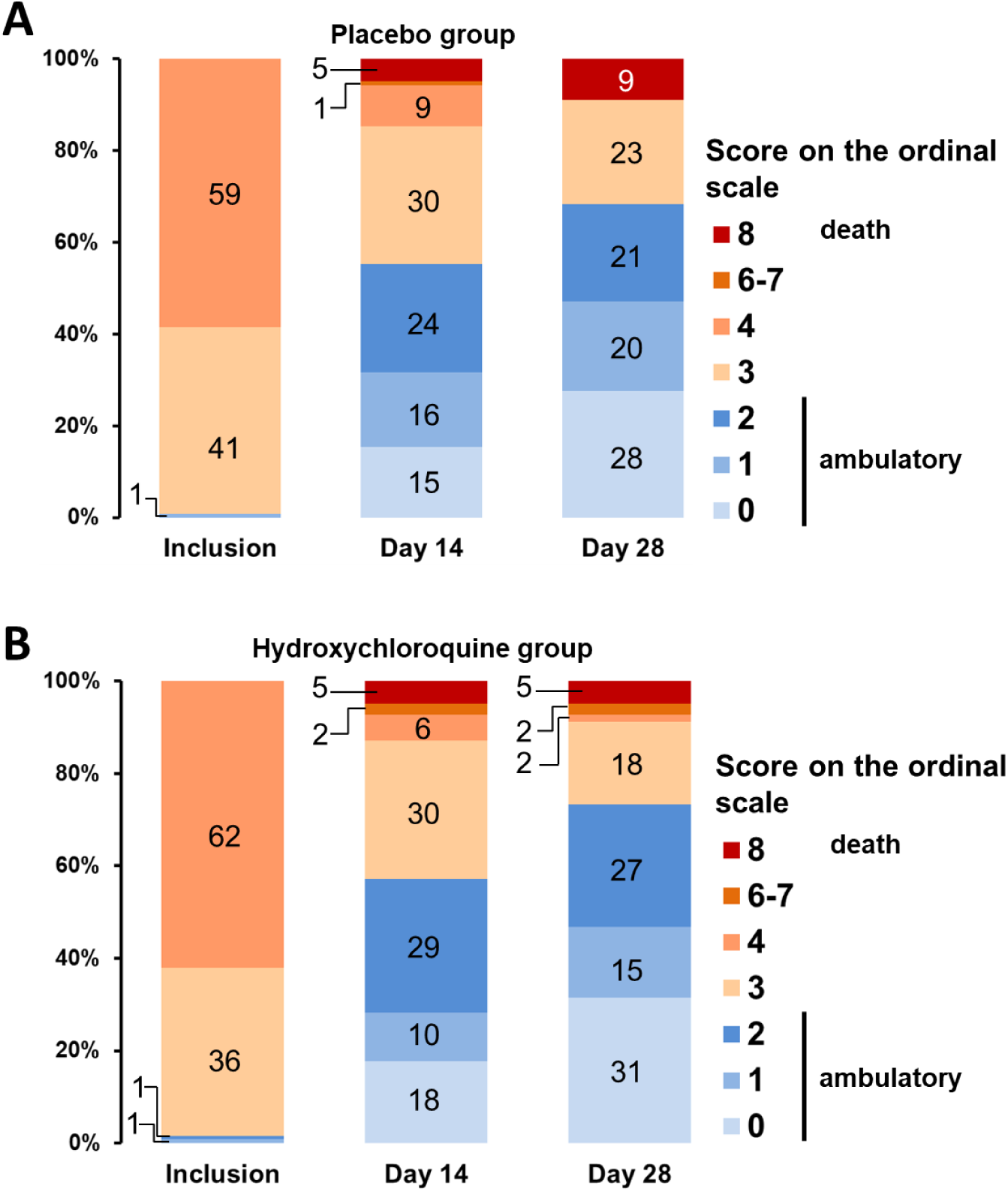
Clinical status at inclusion, Day 14, and Day 28 according to treatment group. Panel A shows the clinical status of patients allocated to the placebo (n = 123) at inclusion, Day 14, and Day 28. Panel B shows the same for patients allocated to hydroxychloroquine (n = 123). Data are missing at Day 14 and 28 for two patients in each group. No patient had a score of 5 (non-invasive ventilation or high-flow oxygen) at Day 14 or 28. The Ordinal Scale for Clinical Improvement is proposed by the World Health Organization as an outcome measure. The score reads as follows: 0: patient uninfected, no clinical or virological signs of infection; 1: patient at home, without limitation of activities; 2: patient at home, with limitation of activities; 3: patient hospitalized without oxygen therapy; 4: patient with oxygen therapy by mask or nasal prongs; 5: patient under non-invasive ventilation or high-flow oxygen; 6: patient under invasive mechanical ventilation; 7; patient under invasive mechanical ventilation and additional organ support, including vasopressors, renal replacement therapy, and extracorporeal membrane oxygenation; 8: death.

Viral shedding could be assessed by RT-PCR in 203 and 174 patients at Day 5 and 10, respectively (Table 2). The rate of SARS-CoV-2-positive RT-PCR tests at Day 5 and 10 was 72.9% (148/203) and 56.9% (99/174), respectively. No significant difference was observed between the two groups, in the global population, as well as in all analyzed subgroups (Table 2 and Figure 3).

### Adverse events

Adverse events occurred in 70 of the 124 patients treated with hydroxychloroquine (56.5%) compared to 61 of the 120 patients who received the placebo (50.8%) (Table 3). Serious adverse events were infrequent and occurred at the same frequency in both groups.

**Table 3.** Adverse events in the safety population according to treatment

| **Event** | **Placebo**<br>**(N = 120)** | **Hydroxychloroquine**<br>**(N = 124)** |
| --- | --- | --- |
| Any adverse event - no. of patients (%) | 61 (50.8) | 70 (56.5%) |
| Hypoglycemia | 2 | 0 |
| Nausea/vomiting | 5 | 7 |
| Vision disorders | 3 | 3 |
| Cardiac rhythm or conduction disorders | 7 | 6 |
| Rash or pruritus | 7 | 10 |
| Diarrhea | 6 | 7 |
| Anemia | 1 | 5 |
| Alanine aminotransferase increased | 1 | 3 |
| Aspartate aminotransferase increased | 1 | 2 |
| Gamma-glutamyltransferase increased | 1 | 3 |
| Headache | 1 | 3 |
| Psychiatric and sleep disorders | 5 | 5 |
| Acute kidney injury | 0 | 4 |
| Hypokalemia | 3 | 1 |
| Vertigo | 2 | 0 |
| Fall | 3 | 1 |
| Dehydration | 1 | 2 |
| Adverse event leading to discontinuation of treatment - no. of patients (%) | 2 (1.7) | 4 (3.2) |
| Cardiac rhythm or conduction disorders | 1 | 4 |
| Skin rash | 1 | 0 |
| Serious adverse event - no. of patients (%) | 4 (3.3) | 3 (2.4) |
| Cardiac rhythm or conduction disorders | 3 | 3 |
| Rash | 1 | 0 |

Only adverse events that occurred in at least two patients are mentioned in the table.

## Discussion

In this trial involving patients with mild-to-moderate COVID-19 at higher risk of worsening, we did not observe a significant difference in the rate of death or tracheal intubation at 14 days following inclusion between patients treated with hydroxychloroquine and those who received the placebo. The rate of serious adverse events was also similar in the two groups.

The strength of this study lies in its double-blind, placebo-controlled design and in the fact that we used consensual strong clinical and virological judgment criteria. Conversely, the lack of power is the main limitation of our trial. Indeed, the study was prematurely stopped after the inclusion of 19% of the planned number of patients due to the slowdown of the epidemic and following the publication of a subsequently retracted study raising concerns over hydroxychloroquine cardiovascular toxicity.

We did not identify any safety concerns related to hydroxychloroquine use. However, we excluded patients with conditions that put them at risk of increased hydroxychloroquine toxicity, such as hypokalemia, prolonged corrected QT interval, or concomitant treatment with an increased risk of torsade de pointes. Of note, hypokalemia has been identified as a frequent complication of COVID-19 [22], and its presence is associated with disease severity. The exclusion of hypokalemic patients may have contributed to the low rate of adverse course that we observed.

The frequency of the primary endpoint, a composite of death or the need for invasive mechanical ventilation at Day 14, was much lower than expected (6.9% versus 20%). This discrepancy may arise from several factors. First, recent data have demonstrated mortality rates lower than those observed in the early phases of the epidemic in Wuhan, on which we based the estimate of the number of needed patients [20, 21]. Second, 15% of the patients included in this study have been treated with corticosteroids during the disease course, and it has been recently demonstrated that dexamethasone decreases mortality in severe COVID-19 [4]. Finally, we excluded patients with an organ failure requiring intensive care or needing more than 3 L/min of oxygen during the initial evaluation. This was based on the hypothesis that antiviral agents are more likely to be effective if they are prescribed early in the course of the disease. On the basis of epidemiological studies, we suspected that COVID-19 patients with advanced age and/or with serious comorbidity would frequently worsen, even if they had no sign of severity at baseline. Our results show that most of them had an uncomplicated course with or without hydroxychloroquine.

Indeed, we did not observe any difference between patients assigned to placebo and those assigned to hydroxychloroquine regarding either mortality or clinical evolution. This is in line with the results observed in randomized open-label studies involving hospitalized patients [10, 14], as well as with a double-blind, placebo-controlled trial evaluating early hydroxychloroquine treatment in ambulatory patients [11]. All these trials involved younger patients with a median age of 40 to 50 years. In our trial, the median time from symptom onset to treatment initiation was 5 days; in the other aforementioned studies, it ranged from 2 to 7 days. Together, the results obtained in these high-quality trials indicate that hydroxychloroquine alone has little chance of having clinical efficacy in COVID-19, regardless of the population considered and the stage of the disease.

Similarly, we observed no benefit of hydroxychloroquine therapy on the duration of RT-PCR positivity. Previous data concerning the impact of hydroxychloroquine on the duration of viral shedding were conflicting. While an uncontrolled study found that the drug alone or combined with azithromycin induced a rapid decrease in the rate of RT-PCR positivity [9], another found a decreased speed of viral clearance [23], and two randomized open-label studies showed no effect [14, 24]. Of note, the decrease in the rate of positive RT-PCR tests observed in our study is in agreement with previous works, which reported a median duration of detectable viral shedding of 14 days after symptom onset in mild COVID-19 and 21 days in severe cases [25-27].

In conclusion, this study failed to detect any effect of hydroxychloroquine on clinical evolution or on the kinetics of viral shedding in patients with mild-to-moderate COVID-19 at higher risk of worsening. Our results do not support the use of hydroxychloroquine alone in this population.

## Supporting information

Study protocol

Statistical Analysis Plan

Members of the HYCOVID study group

## Data Availability

Deindentified data will be made available upon reasonable request to the study data manager.

## Funding

This work was supported by a grant from the French Ministry of Health through a national call for proposals for therapeutic trials on COVID-19.

The trial also received an exceptional donation from the *Pays de la Loire* region and from the *Angers Loire Métropole* conurbation.

## IRB Approval

The Sponsor obtained approval from the *Comité de Protection des Personnes du Sud-Ouest et Outre-Mer 4* (No CPP2020-03-036 / 2020-001271-33 / 20.03.24.72431) and from the *Agence Nationale de Sécurité du Médicament et des produits de santé* (ANSM; No MEDAECNAT-2020-03-00045).

## Acknowledgments

The authors would like to thank all patients who participated to the study, and all people who contributed to the trial. A complete list of investigators and other people implicated in the study is provided in the supplementary appendix.

## Notes

### Competing Interest Statement

The authors have declared no competing interest.

### Clinical Trial

ClinicalTrials.gov Identifier : NCT04325893

### Author Declarations

The Sponsor obtained approval from the Comite de Protection des Personnes du Sud-Ouest et Outre-Mer 4 (No CPP2020-03-036 / 2020-001271-33 / 20.03.24.72431) and from the Agence Nationale de Securite du Medicament et des produits de sante (ANSM; No MEDAECNAT-2020-03-00045).

