## Supplementary material for "A placebo-controlled double blind trial of hydroxychloroquine in mild-to-moderate COVID-19": Study protocol

### A prospective, multicentre, randomised, double-blind, parallel-group, superiority study to assess, in non-severe COVID-19 patients with a high-risk of complication, the efficacy of hydroxychloroquine versus placebo to avoid intubation with mechanical ventilation or death: Hydroxychloroquine versus placebo in COVID-19 patients at risk for severe disease.

### HYCOVID

**Coordinating Researcher:**
Dr. Vincent Dubée
Department of Infectious and Tropical Diseases
CHU d'Angers (Angers University Hospital)
4 rue Larrey
49933 Angers CEDEX 09

**Research Sponsor:**
CHU d'Angers (Angers University Hospital)
**Contact:**
Management and Development Unit
Delegation for Clinical Research and Innovation (DRCI)
CHU d'Angers (Angers University Hospital)
4 rue Larrey
49933 Angers CEDEX 09

**Department Responsible for Data Management:**
Unit for Data Management and Assessment
Delegation for Clinical Research and Innovation (DRCI)
CHU d'Angers (Angers University Hospital)
4 rue Larrey
49933 Angers CEDEX 09

This protocol is the property of Angers University Hospital (CHU d'Angers). It should only be used for conducting this study. The protocol should not be shared with any persons not involved with the study or used for any other purpose without the prior written consent of Angers University Hospital.

*(Pursuant to version 3 dated 12/16/2019 of the framework protocol of Angers University Hospital)*

### SIGNATURE

### Hydroxychloroquine Versus Placebo in Patients Presenting non severe COVID-19 and at Risk of complicated course: A Prospective, Multicenter, Randomized, Double-Blind Study.
### HYCOVID

#### ABSTRACT

**Study Classification**

Interventional study – Drug

**Context and Grounds for Research**

A new human coronavirus, SARS-CoV-2, emerged in China in December 2019 and has spread rapidly. COVID-19, the disease caused by this virus, has a very polymorphous clinical presentation, which ranges from isolated attacks on the upper respiratory tract to acute respiratory distress syndrome. It may appear serious straightaway or may develop in two stages, with a worsening condition 7 to 10 days after the initial clinical signs, potentially linked to a cytokine storm in the immune system and accompanied by a high risk of thrombosis. The global mortality rate of COVID-19 seems between 3% and 4%, the elderly being particularly vulnerable to SARS-CoV-2.

Treatment is largely symptomatic as no treatment to date has shown to be of clinical benefit for this illness. Hydroxychloroquine is a derivative of chloroquine, which is commonly used to treat some autoimmune diseases, such as systemic lupus erythematosus. It is active *in vitro* in cell models of infection by multiple viruses, such as HIV, hepatitis C, and SARS-CoV. However, its efficacy in viral infections among humans has not been demonstrated.

Very recently, a non-controlled preliminary study looked into the efficacy of hydroxychloroquine on the viral shedding of subjects affected by COVID-19. Among 20 patients treated with hydroxychloroquine in doses of 600 mg per day, the percentage of nasopharyngeal swab sample positive for SARS-CoV-2 using RT-PCR went from 100% on inclusion (start of treatment) to 43% six days later. In comparison, among the 16 patients who were not treated, 90% had a positive RT-PCR six days after inclusion. Furthermore, hydroxychloroquine has immunomodulating and anti-inflammatory properties, which could theoretically prevent or limit secondary complications.

The research hypothesis is that treatment with hydroxychloroquine improves prognosis and reduces the risk of death or intubation and invasive ventilation on patients suffering from COVID-19 who initially do not meet the severity criteria, but are at high risk of deteriorating.

**Objectives**

*The primary objective* is to evaluate the efficacy of hydroxychloroquine versus placebo on the rate of mortality and use of invasive ventilation in the 14 days following the start of treatment among non-severe COVID-19 patients at high risk of complicated course.

*The secondary objectives are:*

1) To evaluate the efficacy of hydroxychloroquine versus placebo among COVID-19 patients with regard to:

a- rate of mortality or use of invasive ventilation during the 28 days following inclusion and start of treatment b- clinical improvement using the WHO Ordinal Scale for Clinical Improvement for COVID-19

c- all-cause mortality

d- virus shedding duration

e- incidence of venous thrombo-embolism occurrences

2) To evaluate the efficacy of hydroxychloroquine versus placebo in the subgroup of patients aged 75 years or older with regard to:

a- clinical improvement using the WHO Ordinal Scale for Clinical Improvement for COVID-19

b- all-cause mortality

3) To evaluate the safety of hydroxychloroquine versus placebo on the occurrence of serious adverse events

*Ancillary objectives*

To evaluate within a subgroup of COVID-19 patients the impact of hydroxychloroquine versus placebo on cytokines and biological markers of immunity, inflammation, and hemostasis.

**Evaluation Criteria**

*Primary criteria*:

Death, regardless of cause, or the need for intubation and invasive ventilation within the 14 days (day 14) following inclusion and the start of treatment (day 0).

*Secondary evaluation criteria*:

Death, regardless of cause, or the use of intubation and invasive ventilation at day 28.

1a and 2a) death, regardless of cause, or the use of intubation and invasive ventilation within the 28 days (day 28) following inclusion and the start of treatment (day 0).

1b and 2b) clinical improvement using the WHO Ordinal Scale for Clinical Improvement for COVID-19 between day 0-day 14 and day 0-day 28.

1c and 2c) All-cause mortality at day 14 and day 28.

1d) The rate of RT-PCR tests positive for SARS-CoV-2 on naso-pharyngeal swab samples at day 5 and day 10.

1e) The rate of symptomatic venous thrombo-embolism events at day 14 and day 28, documented and confirmed by an adjudication committee.

*Secondary tolerance criteria*

3) The rate of serious adverse events at day 28, defined in accordance with current regulations.

*Ancillary criteria*

The changes of cytokines and biological markers of immunity, inflammation, and hemostasis between day 0, day 5 and day 10.

**Research Plan and Progression**

HYCOVID is a multicenter, controlled, randomized, double-blind, superiority trial versus placebo in two parallel groups.

Patients with SARS-CoV-2 infection, hospitalized or at home, are assessed for potential inclusion. Patients who meet the inclusion criteria and do not meet the exclusion criteria are included. Randomization is performed using a minimization algorithm based on several criteria (duration of disease, risk factors of adverse event, COVID-19 diagnosis modality, hospitalization, concomitant treatments, center). The treatment batch includes tablets for the nine days of treatment; it is not possible to make a distinction between the hydroxychloroquine and the placebo. The first dose of 400 mg is taken immediately after inclusion at day 0, the second dose of 400 mg is taken on the same evening, and the treatment is then continued for the following eight days at a rate of 200 mg in the morning and evening.

The nasopharyngeal samples are taken to perform RT-PCR SARS-CoV-2 at day 5 and day 10. A clinical and/or telephone follow-up is carried out at day 14 and day 28.

For a subgroup of 200 patients in the context of an ancillary study, samples to perform the dosage of the biological markers are taken at day 0, day 5, and day 10 and frozen for later analysis.

An independent data and safety monitoring board (IDSMB) carries out a follow-up of the safety and efficacy data, supported by the interim analyses of the primary evaluation criteria carried out every 50 patients in order to establish recommendations on the need to stop or to continue the study. If a difference in efficacy is detected early, this will lead to premature termination of the study in order to make active treatment immediately available for COVID-19 patients.

The statistical analyses will be carried out with the intention of treating the entire included and analyzable population.

**Inclusion Criteria**

- Adult patient
- Symptomatic COVID-19 confirmed by RT-PCR SARS-CoV-2 or, failing that, by thoracic CT scan suggesting viral pneumopathy of peripheral predominance in a clinically significant context.
- COVID-19 diagnosed within the two calendar days or, in patients asymptomatic at the time of postive RT-PCR, onset of symptoms within two calendar days
- Patient having at least one of the following risk factors for developing complications:
    - age ≥75 years old
    - Age between 60 and 74 years old and presence of at least one comorbidity among the following: obesity (body mass index ≥ 30 kg/m$^2$), arterial hypertension requiring treatment, diabetes mellitus requiring treatment
    - oxygen dependence characterized, in the 24 hours preceding the inclusion, by peripheral capillary oxygen saturation less than or equal to 94% ($SpO2 \leq 94\%$) in ambient air, or a ratio of partial oxygen pressure to fraction of inspired oxygen less than or equal to 300 mmHg ($PaO2/FiO2 \leq 300$ mmHg).
- Electrocardiogram showing absence of corrected QT prolongation greater than 440 ms in men and 460 ms in women.
- Patient affiliated with or beneficiary of a social security scheme.
- Written and signed consent of the patient or their representative or, if not possible, emergency inclusion procedure.

**Exclusion Criteria**

- Last RT-PCR negative for SARS-CoV-2
- Peripheral capillary oxygen saturation less than or equal to 94% ($SpO_2$ ≤94%) despite oxygen therapy being greater than 3 L/min (> 3 L/min)
- Organ failure requiring admission to a critical or intensive care unit.
- Porphyria, known deficit of glucose-6-phosphate dehydrogenase
- Comorbidity that is life threatening in the short-term (life expectancy < 3 months)
- Any reason that makes patient follow-up throughout the study impossible
- Current treatment with hydroxychloroquine
- Absolute contraindication to treatment with hydroxychloroquine (known hypersensitivity, concomitant treatment with risk of torsades de pointe)
- Hypokalemia of less than 3.5 mmol/L
- Corrected QT prolongation higher than 440 ms in men and 460 ms in women.
- Child C-class liver cirrhosis
- Chronic kidney failure with estimated GFR of ≤ 30 ml/min (for patients treated with azithromycin at time of inclusion, eGFR ≤ 40 ml/min)
- Women who are pregnant, breastfeeding, or parturient

**Data Collection and Circulation**

The data are collected in an electronic CRF (Ennov Clinical). Patients are identified by the sequence number of their inclusion in the study.

The database conforms to the recommendations set by the French Data Protection Authority (CNIL) (MR01) in terms of identifiable data (no surname, first name, date of birth recorded as month/year only, initials of surname and first name recorded), data management, and data security.

In centers that do not have sufficient staff to carry out the telephone follow-up at day 14 and day 28, centralized telephone follow-up will be set up subject to the authorization of CNIL.

**Number of People Participating in the Study**

Based on data from the literature, the rate of patients requiring respiratory support and/or who dead is estimated at 20% in the study population. A headcount of 615 patients per group must be studied to demonstrate, under a bilateral hypothesis, an absolute difference of 6% between the two groups (relative difference of 30%) with an alpha risk of 5% and 80% power.

Taking into account patients lost to follow-up and those who cannot be evaluated (estimated at 5%), it is necessary to include **1,300 patients** in total.

**Research Duration**

Duration of inclusion: 4 months maximum

Duration of participation: 28 days

Study duration: 5 months maximum

**Expected Outcomes and Perspectives**

The expected benefit of hydroxychloroquine is significant. The expected effect is an absolute decrease in the requirement for invasive ventilation and/or in death estimated at 6%. COVID-19 could affect more than half of the French population in the coming

| |
|---|
| weeks. This condition is particularly serious in the targeted population: older people and patients who have lung damage with oxygen dependency.<br><br>The possibility of carrying out this double-blind study in an extremely short time due to the immediate availability of the treatment and its specific placebo, its multicentric nature allowing the inclusion of the necessary patients in a few weeks, and the possibility of stopping the study early if the interim analyses sent to the IDSMB demonstrate the superiority of hydroxychloroquine compared to the placebo, will allow the results to be available very quickly, benefiting all COVID-19 patients in France and internationally. |

#### SUMMARY

### 1. Context and Scientific Grounds for Research

#### 1.1. COVID-19

A new human coronavirus that is transmitted through respiratory droplets, SARS-CoV-2, emerged in China in December 2019 and has spread rapidly across the Northern Hemisphere (1). COVID-19, the disease caused by this virus, has a very polymorphous clinical presentation, which ranges from isolated attacks on the upper respiratory tract to acute respiratory distress syndrome and includes digestive and neurological signs and symptoms. It may appear serious straightaway or may develop in two stages, with a worsening condition 7 to 10 days after the initial clinical signs, potentially linked to a cytokine storm in the immune system (2). Immunological analyses of patients suffering from a severe form have shown a reduction in lymphocytes, particularly $CD4^+$ T cells, and an increase in the rate of inflammatory cytokines (TNF-α, IL-1 and IL-6) (2). Patients suffering from severe causes of COVID-19 also present a state of thrombophilia, as shown by an increase in D-dimers and a higher risk of pulmonary embolism (3).

The global mortality rate of COVID-19 is understood to be between 3% and 4%, with the more severe forms being more frequent among older age groups (4). Recent data indicate that the risk of unfavourable evolution is higher in patients with arterial hypertension (5), diabetes mellitus (6), or obesity (7).

Treatment is largely symptomatic as no antiviral treatment to date has shown to be of clinical benefit for this illness (8). However, several compounds have been identified as being *in vitro* inhibitors of the virus (9). Of these compounds, chloroquine and hydroxychloroquine are currently receiving an important media attention.

#### 1.2. Hydroxychloroquine and Chloroquine

Hydroxychloroquine (HCQ) and chloroquine (CQ) are synthetic antimalarials that are derivatives of quinine, taken orally, and part of the 4-aminoquinoline group (10). The HCQ derivative of chloroquine has been manufactured since 1950 and is distinguished from chloroquine by the presence of a hydroxyl on an ethyl group.

The therapeutic properties of these two drugs go far beyond their antimalarial purposes. In fact, they are now used as a common treatment for certain inflammatory diseases, such as systemic lupus erythematosus and rheumatoid arthritis. They are also prescribed in the treatment of some chronic infections caused by intercellular bacteria (Whipple disease and Q fever). Lastly, their anticancer properties are also well documented, even if they are not used in oncology (11).

*Anti-inflammatory properties*

Synthetic antimalarials have been used since the 1950s for systemic lupus erythematosus (12) and now constitute a cornerstone of the basic treatment for this disease. HCQ is also widely used against other autoimmune diseases: rheumatoid arthritis, Sjögren’s syndrome, and thrombotic antiphospholipid syndrome (13).

The mechanisms that underpin the immunomodulating effect of HCQ and CQ are complex. These include weak bases that alkalinize acidic vesicles, particularly lysosomes (14). Modifying the pH of the lysosome changes the function of numerous enzymes, which will have an effect on, in particular, processing antigens (15) and on the maturation of proteins involved in the immune system. HCQ and CQ inhibit the production of pro-inflammatory cytokines, including IL1, IL6, MIP-1ß, TNFα, IFNα, and IFNγ (16).

*Anti-thrombotic action*

HCQ is effective in preventing venous thrombo-embolism occurrences in patients suffering from antiphospholipid syndrome (13) because of a mechanism that is little understood, but probably goes beyond its anti-inflammatory effect (17). It reduces platelet activation and aggregation in several experimental *in vitro* and animal models. It also reduced blood viscosity (18).

#### 1.3. Hydroxychloroquine and Viral Infections

*Antiviral action of HCQ and CQ*

The discovery of CQ's antiviral effect dates back to the end of the 1960s (19). CQ and HCQ are active in cellular models of infections caused by several viruses, including the coronaviruses OC43 and SARS-CoV, enterovirus A71, the Zika virus, H5N1 influenza virus, HIV, the CHIKV virus that causes Chikungunya, hepatitis C, and the Ebola virus. There are likely many mechanisms that underpin this antiviral effect. The increase in the pH of endosomes caused by HCQ and CQ is potentially what largely explains the effects observed, as the activity of certain viral enzymes varies depending on the pH.

In animal models, the activity of CQ and HCQ is proven to be sometimes moderate and most often nonexistent. Similarly, attempts to use CQ and HCQ as antiviral drugs in humans have not revealed significant efficacy. Administration of chloroquine (500 mg/day for one week, then 500 mg/week for 11 weeks) has not demonstrated efficacy in preventing seasonal flu (20). In the case of Chikungunya, despite satisfactory efficacy *in vitro*, hydroxychloroquine has no effect on the clinical presentation of the primary infection, but does increase the risk of developing chronic joint pain (21). In cases of HIV virus infections, the use of CQ has not been proven. The molecule may even strengthen the infectiousness of the virus with regard to the astrocyte reservoir (22). The most interesting results from the field of viral infections were found when treating patients suffering from hepatitis C who did not respond to interferon treatment (23). However, with the appearance of antiviral drugs that have a direct impact against HCV, this line of research was abandoned.

*In vitro activity of HCQ and CQ against SARS-CoV-2*

Chloroquine is active in cellular models of infection with SARS-CoV, which is the closest human virus to SARS-CoV-2 in terms of phylogenetics (24). The mechanism of the antiviral action may be based in the inhibition of the glycosylation terminal of the virus entry point, ACE2.

It is also active in weak concentration in a model of infection with SARS-CoV-2 in Vero E6 cells (25); EC90 is at 6.9 µM, which is a concentration lower than that observed in the plasma of patients treated with chloroquine for rheumatoid arthritis. The molecule appears to be active in the early (adding the molecule before exposure to the virus) and late (adding the molecule after the virus) stages of infection. The same team has shown that hydroxychloroquine is also active in this model, albeit with a slightly lower selectivity index, which suggests that it is slightly less active than chloroquine (26).

*Using HCQ and CQ against COVID-19*

Based on the results of *in vitro* studies, the team at the Institut Hospitalo-Universitaire Méditerranée Infection started a non-controlled, open clinical trial (27). This study aimed to evaluate the impact of treatment with hydroxychloroquine in doses of 600 mg per day on the presence of SARS-CoV-2 in nasopharyngeal samples from 20 patients suffering from COVID-19, including two asymptomatic patients and six suffering from non-serious pneumonia. The results of tests on the virus, which are carried out daily, were compared with those taken from 16 patients who were not treated and were cared for in another institute. The non-treated patients were slightly older (average age of 37 years old in comparison to 51 years old for the treated patients, p = 0.06). From the third day of treatment, there was a significant difference between the two groups, as more than 90% of the non-treated patients tested positive for the virus versus 50% of the patients treated with hydroxychloroquine (p = 0.005). On the sixth day of treatment, 90% of the non-treated patients tested positive for the virus versus 30% of the patients treated with hydroxychloroquine (p = 0.001). Six out of 20 patients treated with hydroxychloroquine also received azithromycin (500 mg on the first day of treatment, then 250 mg/day). Among these patients, the efficacy of treatment appeared more noticeable as the virus could not be detected in any of them from the fifth day of treatment, compared to 7 out of 14 patients treated with hydroxychloroquine only and 3 out of 16 patients who were not treated.

Despite its preliminary nature, this study has been widely broadcast and many doctors have started to use hydroxychloroquine in both France and the rest of the world. There are in fact numerous theoretical arguments in favor of using this molecule: satisfactory *in vitro* antiviral activity, immunomodulating effect that can have a positive impact in patients with a deregulated immune response, and potential anti-thrombotic activity, although incidences of thrombo-embolism are increasingly recognized as contributing to the severity of the disease.

However, it must be noted that the sole sign of this molecule's efficacy in humans is a virological criterion, the clinical relevance of which is still to be proven. The impact of negativation virus shedding on the clinical development of the disease is not known. The connection between the viral load on nasopharyngeal samples and the severity of the disease has been studied in 76 patients, 30 of whom presented a serious form as defined by one of the following factors: respiratory rate ≥ 30/min, resting SpO2 ≤ 93%, PaO2/FiO2 ≤ 300 mmHg, or serious complications of the disease (at least one organ failure) (28). Patients suffering from a serious form of COVID-19 had a viral load at the time of treatment that was, on average, 60 times higher than that presented by benign forms. In this latter group, viral excretion could no longer be detected in the ten days following the appearance of symptoms in 90% of cases. On the other hand, all serious cases had a viral load that could be detected ten days after the appearance of symptoms. These results suggest a positive correlation between initial viral load and severity, as was shown by SARS in 2003-2004 (29).

Another study mentioned the use of CQ against COVID-19 in China (30). Approximately 100 patients were treated using chloroquine sulfate, which had higher efficacy than the control on the clinical and radiological development of the disease. However, the exact data that this article references has not been published. Based on these results, a consensus reached by Chinese experts recommends the use of chloroquine phosphate in doses of 500 mg twice per day to treat COVID-19 (31). Furthermore, the Dutch Centre for Infectious Disease Control and the Italian Society for Infectious and Tropical Diseases advocate the use of CQ or HCQ for patients suffering from COVID-19, while highlighting the low level of evidence to support this recommendation (32). 23 clinical trials using HCQ or CQ against COVID-19 were ongoing in China on 03/01/2020 (33).

#### 1.4. Toxicity of Hydroxychloroquine

The toxicity of HCQ is largely related to ophthalmological toxicity. This manifests a rare variety of maculopathy known as “Bull’s eye” in 0.38% to 1% of patients treated and is largely dependent on the daily dose administered (> 5 or > 6.5 mg/kg/day depending on the study) and the duration of exposure (> 5 years) (33). The risk of macular toxicity in short-term treatments (several days) would be zero.

Muscular and neurological toxic effect have also been reported in relation to HCQ, but remain rare and linked to extended treatments (several years) (34–36). To our knowledge, there is no precise data on the toxicity of HCQ for older patients. Nevertheless, there appears to be indications that are more favorable for conditions specific to older patients: HCQ seems to reduce cardiovascular morbidity-mortality in rheumatoid arthritis (37–39) and reduce the risk of cancer and all-cause mortality in cases of autoimmune diseases (40).

Lastly, HCQ has been the focus of a considerable volume of literature on the risk of cardiomyopathy that highlights two main points. The first concerns the risk of hypertrophic or restrictive cardiomyopathy and is limited to extended and very extended treatments, even if some cases can be connected to significant voluntary ingestion (41). The second toxic event concerns corrected QT interval prolongation: synthetic antimalarials are listed among the treatments at risk of lengthening the QT interval. However, the reported cases of QT lengthening and torsades de pointe involve prolonged treatments based on HCQ and often pathological (hypokalemia) and therapeutic (beta blockers) associations (42–45). Several studies that have been conducted on patients treated with HCQ for autoimmune diseases have not demonstrated a significant QTc interval prolongation (46,47). Lastly, a 2013 pharmacological study by Pfizer laboratories on behalf of the FDA showed that QTc interval prolongation in 116 healthy subjects during co-prescription of high doses of chloroquine (1000 mg/day) and azithromycin were 5 ms, 7 ms, and 9 ms, respectively, for azithromycin in daily doses of 500 mg, 1000 mg, and 1500 mg (48).

HCQ passes the placental barrier in pregnant women, but no malformative effects have been identified (49). It is strongly recommended that patients who are treated with HCQ for lupus continue taking this drug when pregnant (50). In addition, this molecule is indicated in cases of recurring fetal death in patients who are suffering from antiphospholipid syndrome (51).

#### 1.5. Choice of Hydroxychloroquine Dosage

The choice of hydroxychloroquine dosage in the treatment of COVID must take into account the data for *in vitro* activity (EC50) and pharmacokinetic data (52). On the basis of these factors, the following plan has been recommended by the REACTing network:

- 800 mg loading dose at day 0
- then a daily dose of 400 mg/day in two oral administrations

These doses have been approved by the Haut Conseil de Santé Publique (French High Council for Public Health) (https://www.hcsp.fr/Explore.cgi/Telecharger?NomFichier=hcspa20200323_coronsarscovrecomthrap.pdf).

#### 1.6. Research Hypothesis

Our primary research hypothesis is that treatment with hydroxychloroquine improves prognosis and reduces the risk of death or intubation and invasive ventilation on patients suffering from COVID-19 who initially do not meet the severity criteria, but are at high risk of deteriorating.

We also believe that hydroxychloroquine can reduce the risk of venous thrombo-embolic events and is not connected to notable side effects in short courses of treatment.

### 2. Study Objectives and Evaluation Criteria

#### 2.1. Primary Objective and Evaluation Criteria

The primary objective is to evaluate the efficacy of hydroxychloroquine versus placebo on the rate of mortality and use of invasive ventilation among among non-severe COVID-19 patients at a high risk of complicated course.

The primary outcome is death, regardless of cause, or the use of intubation and invasive ventilation within the 14 days (day 14) following inclusion and start of treatment (day 0).

#### 2.2. Secondary Objectives and Evaluation Criteria

##### *2.2.1. Efficacy criteria in the sample group*

The objective is to evaluate the efficacy of hydroxychloroquine versus placebo among COVID-19 patients based on several secondary evaluation criteria:

a- the rate of mortality or use of invasive ventilation within the 28 days (day 28) following inclusion and the start of treatment,

b- clinical improvement using the WHO Ordinal Scale for Clinical Improvement for COVID-19 at day 14 and day 28 (53),

c- all-cause mortality at day 14 and day 28,

d- virus shedding assessed by the rate of RT-PCR positive for SARS-CoV-2 with naso-pharyngeal swab samples at day 5 and day 10,

e- the rate of symptomatic venous thrombo-embolism events (VTE) at day 14 and day 28. VTE should be documented and confirmed by the adjudication committee.

##### *2.2.2. Efficacy criteria in the subgroup of people aged 75 years or older*

The objective is to evaluate the efficacy of hydroxychloroquine versus placebo in the subgroup of people aged 75 years or older with regard to:

a- clinical improvement using the WHO Ordinal Scale for Clinical Improvement for COVID-19 at day 14 and day 28

b- all-cause mortality at day 14 and day 28.

##### *2.2.3. Safety criteria*

The objective is to evaluate the safety of hydroxychloroquine versus placebo on the occurrence of serious adverse events at day 28. Serious adverse events will be evaluated and their provenance will be analyzed in line with the applicable regulations (cf. safety chapter).

#### 2.3. Objectives and Evaluation Criteria of the Ancillary Study (Optional)

The general objective of this ancillary study, which will be carried out on a subgroup of 200 patients treated with either hydroxychloroquine or a placebo, is to build up a collection of standard biological data and a biobank (plasma bank, serum bank, and DNA bank) so as to enable analysis of physiopathological mechanisms.

More specifically, the primary objective is to evaluate the impact of hydroxychloroquine versus placebo on cytokines and biological markers of immunity, inflammation, and hemostasis within a subgroup of COVID-19 patients.

The secondary objective is to evaluate the pharmacokinetics of hydroxychloroquine in COVID-19 patients.

The evaluation criteria of the ancillary study correspond to the change of cytokines and biological markers of immunity, inflammation, and hemostasis between day 0, day 5, and day 10.

The markers that will be analyzed, if possible, are as follows:

- Soluble factors of the innate immune system: cytokines and inflammatory chemokines (IL6, IL1b, TNFa, CCL2, CCL3, CXCL10, G-CSF, and GM-CSF) and type-I IFN
- Activation markers and effector functions of T cells (IFNg, IL2, sCD25, IL-7, IL-17, IL-10)
- Markers of cell death connected to excessive inflammation (circulating tumor DNA) and, more specifically, to neutrophils (MPO)
- Activation markers of thrombosis: D-dimers, thrombin, antithrombin, fibrin monomers, circulating microvesicles, and the fibrin profile of these microvesicles (Prof. Pierre Morange, Prof. Françoise Dignat-George, Marseille) with
  - Luminex assay of four endothelial molecules (sICAM-1, sVACM61, sEsel, and sPsel), making it possible to define endothelial activation
  - Analysis of a Willebrand antigen factor in an ELISA
  - Analysis using an ELISA kit for soluble CD146, a molecule of the endothelial junction that is increased in inflammatory vascular diseases and is a good reflection of vessel permeability and inflammation.
- Hydroxychloroquine concentration

These markers will be analyzed at day 0, day 5, and day 10 to measure the kinetics of the innate immune system after treatment and from a longitudinal perspective. This information will make it possible to associate the beneficial effects of treatment with modulations of the immune system and on hemostasis. The anticipated results are a reduction in the cytokine storm and prothrombotic factors with treatment using hydroxychloroquine.

#### 2.4.Measurement Methods

##### *2.4.1. WHO Ordinal Scale for Clinical Improvement for COVID-19*

The Ordinal Scale for Clinical Improvement (OSCI) is nine-level scale created by the World Health Organization specifically for COVID-19 (53).

0: Patient at home with no clinical or virological evidence of infection
1: Ambulatory patient at home with no limitation of activities
2: Ambulatory patient at home with limitation of activities (or oxygen)
3: Patient hospitalized with mild disease with no oxygen therapy
4: Patient hospitalized with mild disease with oxygen therapy (nasal prongs, mask)

5: Patient hospitalized with severe disease and non-invasive ventilation or high-flow oxygen
6: Patient hospitalized with severe disease, intubation and mechanical ventilation
7: Patient hospitalized with severe disease and intubated with ventilation and additional organ failure requiring invasive treatment (renal replacement therapy, vasopressive amines, or ECMO)
8: Death

##### *2.4.2. Viral shedding by RT-PCR SARS-CoV-2*

Samples specifically for the study are taken with nasopharyngeal swab at day 5 and day 10 during a consultation carried out in the patient follow-up. These samples are taken only for patients whose COVID-19 diagnosis was confirmed by RT-PCR.

The nasopharyngeal swab samples are highly infectious and must be handled with precautions (class II biosafety cabinet).

If possible, the samples will be analyzed in the center where the patient was included. If this is not possible, the samples will be sent to the study sponsor's center (Angers University Hospital (CHU d'Angers)). In this case, each center shall decide where the aliquots of this sample should be carried out (virology laboratory or center for biological resources). Once frozen at -80°C, the sample tubes will be transferred to the Biological Resource Center (BRC) at Angers University Hospital at the end of the study (transport with triple packaging and dry ice) in accordance with current regulations.

##### *2.4.3. Venous thrombo-embolism*

Venous thrombo-embolism (VTE) taken into account in this respect are those that are diagnosed during the follow-up period of 28 days and based on the data in a discharge report, a report of an additional examination, and/or on information provided by general practitioners and patients.

The following events are considered as being venous thrombo-embolism occurrences:

- symptomatic distal or proximal deep vein thrombosis objectively confirmed by a positive venous Doppler ultrasonography, a phleboscanography, or a venography.

- symptomatic pulmonary embolism confirmed by a thoracic computed tomography scan angiography (CTPA), a high-probability V/Q lung scintigraphy, a thoracic MRA, or a pulmonary angiogram or through the combination of a proximal deep vein thrombosis on venous ultrasonography and thoracic symptomatology suggestive of pulmonary embolism (acute dyspnea, chest pain, haemoptysis).

- death caused by incidental pulmonary embolism diagnosed at autopsy

- unexplained sudden death, in which a thromboembolic cause cannot be excluded

An independent adjudication committee evaluates and rules on clinical venous thrombo-embolism occurrences while unaware of the group to which the subject was assigned.

##### *2.4.4. Ancillary study*

The evolution of cytokines and biological markers of immunity, inflammation, and hemostasis is studied as part of an ancillary study that focuses on 200 patients in some volunteering centers that have a Biological Resource Center.

### 3. Study Design

#### 3.1. Study Type

HYCOVID is a multicenter, controlled, randomized, double-blind superiority trial against placebo in two parallel groups.

#### 3.2. Justification of the Number of People Included in the Study

Based on data from COVID-19 in China, the rate of patients requiring respiratory support and/or who dead is estimated at 20% in the study population (high-risk patients). A headcount of 615 patients per group must be studied to demonstrate, under a bilateral hypothesis, an absolute difference of 6% between the two groups (relative difference of 30%) with an alpha risk of 5% and a power of 80%. Taking into account patients lost to follow-up and those who cannot be evaluated (estimated at 5%), it is necessary to include 1,300 patients in total.

#### 3.3. Randomization

Randomization is carried out immediately after the patient is included.

Randomization was performed using a web-based system (Ennov Clinical® software) using a minimization algorithm (dynamic randomization) based on eight criteria:

- Situation at risk for unfavourable evolution:
  - age older than 75 years
  - oxygen requirement
  - age older than 75 years and oxygen requirement
  - age between 60 and 74 years old and presence of comorbidities
  - age between 60 and 74 years old and presence of comorbidities and oxygen requirement
- diagnostic criteria of positive COVID-19 (RT-PCR or scan)
- first symptoms of COVID-19 dating back less than seven days (yes/no)
- hospitalization (yes/no)
- concomitant treatment with azithromycin (yes/no)
- concomitant treatment with antiviral drug (yes/no)
- treatment with corticosteroids (yes/no)
- center

The randomization process was conducted by the Biostatistics and Methodology Department at Angers University Hospital.

The patients are randomized using Ennov Clinical® randomization software and before receiving the corresponding treatment.

#### 3.4. Blinding

##### *3.4.1. Methods for implementing and maintaining blinding*

This study is double-blind with the use of a placebo that is impossible to distinguish from hydroxychloroquine tablets by appearance, odor, or taste (cf. chapter 5). The presentation of treatments does not allow a distinction between HCQ and the placebo. Each item of treatment has a unique processing number. Only this number can identify the substance. The link between each processing number and the content is only known by the coordinating pharmacy at Angers University Hospital and the Creapharm Group pharmaceutical institute.

The coordinating pharmacy at Angers University Hospital supervises the production, packaging, and distribution of treatment batches (HCQ/placebo). These tasks are carried out by an external service provider (Creapharm Group pharmaceutical institute). A contract binds the sponsor to the external service provider.

Furthermore, the doctors and nurses taking care of the patients, as well as the patients themselves, do not know which treatment they have been assigned. The study personnel, including the person responsible for centralized telephone meetings, are also blind.

##### *3.4.2. Indications and methods of unblinding*

Indications of Unblinding

Unblinding by the researcher during a clinical trial should only take place in exceptional circumstances. Unblinding may be undertaken by the researcher in the following circumstances:
- When knowledge of the investigational product received is needed to care for the person participating in the trial, especially in the event of a serious adverse event, and medical care differs depending on the treatment received
- In the event of a death that is unexplained or potentially linked to the investigational product
- In the event that it is accidentally or intentionally taken by an individual who is not the person participating in the trial
- Or in any other event that jeopardizes the safety of the person participating in the trial

If unblinding is carried out under conditions other than those described above, a protocol deviation will be noted. Angers University Hospital will implement corrective and preventative actions within the center.

Methods of Unblinding

Unblinding takes place using a secure electronic system (Ennov Clinical software). The procedure is given in the eCRF user guide.

In the event that the electronic system does not work, a downgraded process using paper forms is available. Unblinding will also be carried out the Angers University Hospital coordinating pharmacy.

This situation may occur in exceptional cases, such as:

- In the event of a serious adverse event, it is vital to know the type of treatment received as the medical care to be given may differ depending on the treatment administered
- In the event of an unexplained death
- In the event that the drug is accidentally ingested by another person
- In any other situation that compromises the health and safety of participants

In any situation not listed above, the unblinding process will be the subject of a protocol deviation notification.

### 4. Selecting Research Subjects

#### 4.1. Inclusion Criteria for Study Participants

- Age ≥ 18 years old

- Symptomatic COVID-19 confirmed by RT-PCR SARS-CoV-2 or, failing that, by thoracic CT scan suggesting viral pneumopathy of peripheral predominance in a clinically significant context.

- COVID-19 diagnosed within the two calendar days or, in patients asymptomatic at the time of postive RT-PCR, onset of symptoms within two calendar days

- Patient having at least one of the following risk factors for developing complications:

    - age ≥75 years old

    - Age between 60 and 74 years old and presence of at least one comorbidity among the following: obesity (body mass index ≥ 30 kg/m$^2$), arterial hypertension requiring treatment, diabetes mellitus requiring treatment

    - oxygen dependence characterized, in the 24 hours preceding the inclusion, by peripheral capillary oxygen saturation less than or equal to 94% (SpO2 ≤ 94%) in ambient air, or a ratio of partial oxygen pressure to fraction of inspired oxygen less than or equal to 300 mmHg (PaO2/FiO2 ≤ 300 mmHg). The following formula is used for this: FiO2 = 21% + 3% per L/min of oxygen therapy (Murray and Nadel's Textbook of Respiratory Medicine, 6th Edition, 2016).

- Electrocardiogram showing absence of corrected QT prolongation greater than 440 ms in men and 460 ms in women.

- Patient affiliated with or beneficiary of a social security scheme.

- Written and signed consent of the patient or their representative or, if not possible, emergency inclusion procedure.

#### 4.2. Exclusion Criteria for Study Participants

- Last RT-PCR negative for SARS-CoV-2

- Peripheral capillary oxygen saturation less than or equal to 94% (SpO2 ≤94%) despite oxygen therapy being greater than 3 L/min (> 3 L/min)

- Organ failure requiring admission to a critical or intensive care unit.

- Porphyria, known deficit of glucose-6-phosphate dehydrogenase

- Comorbidity that is life threatening in the short-term (life expectancy < 3 months)

- Any reason that makes patient follow-up throughout the study impossible

- Current treatment with hydroxychloroquine

- Absolute contraindication to treatment with hydroxychloroquine (known hypersensitivity, retinopathy, concomitant treatment with risk of ventricular disorders, particularly torsades de pointe*)

- Hypokalemia of less than 3.5 mmol/L

- Corrected QT prolongation higher than 440 ms in men and 460 ms in women.

- Child C-class liver cirrhosis

- Chronic kidney failure with estimated GFR of ≤ 30 ml/min (for patients treated with azithromycin at time of inclusion, eGFR ≤ 40 ml/min)

- Women who are pregnant**, breastfeeding, or parturient

** citalopram or escitalopram (Seroplex®, Seropram®), hydroxyzine (Atarax®), domperidone (Motilium®, Peridys®), piperaquine (Eurartesim®), amiodarone (Cordarone®), flecainide (Flecaine®), mexiletine.*

*** Women of reproductive age must agree to have a blood pregnancy test (unless this has already been done prior as part of treatment or if pregnancy is ruled out).*

### 5. Care and Treatment Administered to Persons Participating in the Study

#### 5.1. Investigational Products

The drugs used over the course of the study are Plaquenil® 200 mg for the treatment group and the placebo for the control group.

A pharmaceutical index indicating all methods related to providing, storing, administering, and managing stocks and to the traceability of investigational products in the centers is established and sent to all the centers with the standard administrative documents.

##### *5.1.1. Study treatment: Plaquenil® 200 mg*

The investigational product 1 (IP 1) is Plaquenil® 200 mg (hydroxychloroquine). It is given in the form of a coated tablet containing 200 mg of hydroxychloroquine sulfate. The excipients are as follows: lactose monohydrate, povidone, starch, magnesium stearate, and, for the coating: hypromellose, macrogol 4000, titanium dioxide (E171), lactose monohydrate. It is administered orally.

The manufacturing laboratory is Sanofi-Aventis France (13 boulevard Romain Rolland, 75014 Paris, France). The marketing authorization number for Plaquenil® is: 364-414-6.

##### *5.1.2. Comparison treatment: Placebo*

The investigational product 2 (IP 2) is the placebo for Plaquenil® 200 mg. It is given as a tablet identical to the Plaquenil® (IP 1) tablet in order to maintain the double-blind aspect. The excipients are as follows: lactose monohydrate, microcrystalline cellulose, anhydrous colloidal silica, magnesium stearate, and, for the coating: Opadry II HP85F18422 white. It is administered orally. The manufacturing pharmaceutical institute is Unither Développement Bordeaux (Z.A. Tech Espace Bat 16, 33185 Le Haillan, France).

##### *5.1.3. Manufacturing of treatments*

The investigational products are manufactured in accordance with good manufacturing practices (GMP).

##### *5.1.4. Packaging and labeling*

The primary packaging in a blister packet of ten tablets and blinding of investigational products are carried out by Creapharm in compliance with applicable regulations.

The secondary packaging in a box of 20 tablets (two blister packets of ten tablets) is carried out by Angers University Hospital's coordinating IHP. This packaging corresponds to nine days of treatment for one patient.

The labeling is carried out by Creapharm and by the coordinating IHP at Angers University Hospital in compliance with the decree of May 26, 2006 establishing the content of labels for investigational products. The treatments are numbered according to the sequence list created by the Unit for Data Management and Assessment at Angers University Hospital. This treatment number is given on the labels of boxes and blister packs.

During randomization, the Ennov Clinical software assigns each patient a treatment number that matches the group assigned in the minimization method (one unique treatment number per box of 20 Plaquenil® 200 mg or placebo tablets).

A counter label is generated by the coordinating IHP at Angers University Hospital in compliance with the decree of May 26, 2006. This counter label states:

- Name of the study
- EudraCT number
- The packaging of two blister packets of ten tablets in a box.
- Name of the research coordinator

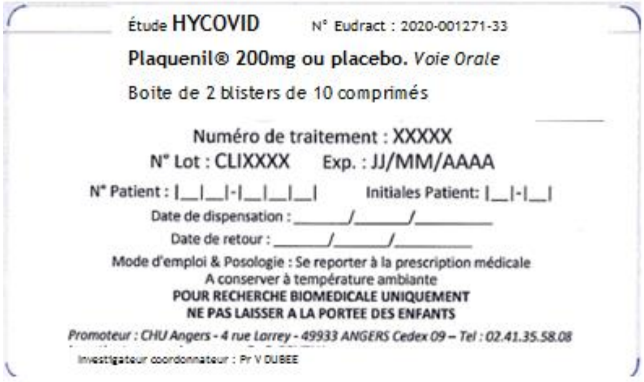

Secondary packaging label (box)

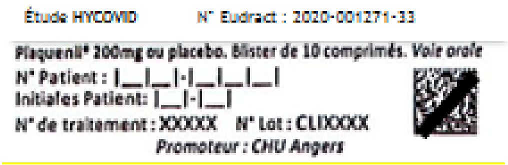

Primary packaging label (blister packet)

##### *5.1.5. Distribution of treatments*

The distribution of investigational products in research centers is carried out by the coordinating IHP at Angers University Hospital. Products are supplied based on the inclusion rates.

##### *5.1.6. Administration*

The investigational products are administered orally. The dosage is four tablets on the first day in two stages (with an interval of at least 4 hours) and then two tablets per day in two stages (one tablet in the morning and evening) over eight days. The tablets should not be crushed, broken, or chewed before oral administration, except for administration through an enteral feeding tube (see annex 2).

The drug is taken after a meal.

The total duration of the treatment is nine days.

##### *5.1.7. Adjustment of the dosage*

There is no adjustment of the dosage.

##### 5.1.8. *Precautions for use*

The precautions for using Plaquenil® are given in the Summary of Product Characteristics (SPC (French only): http://agence-prd.ansm.sante.fr/php/ecodex/index.php).

##### 5.1.9. *Conditions for storing investigational products*

The investigational products are stored at room temperature.

##### 5.1.10. *Follow-up strategy for monitoring investigational products*

The patient's initials and number are provided on the labels (box and blister packet) when dispensing the treatment.

Once used, the boxes containing empty, full, or partially used blister packets are kept and returned to the center's pharmacy.

Throughout the study and after the treatments are checked by the clinical research associate (CRA), the returned treatments may be destroyed on site, ONLY after written authorization from the sponsor (coordinating IHP).

##### 5.1.11. *Management of stocks, resupplying, and dispensing treatment*

The procedures used to deliver, supply, and trace the products at the centers are described in a pharmaceutical appendix that was submitted to the researchers and the pharmacists of the IHP in every site during the inclusion visits.

The first supply of stock and replenishment of stock is managed automatically for pharmacies at each center. They are managed by the coordinating IHP at Angers University Hospital.

Resupply is based on the inclusion rates.

The treatments are dispensed by the pharmacy at each research center in accordance with the applicable regulations on clinical trials.

The treatment number is assigned to the patient using Ennov Clinical software during their inclusion on day 0.

##### 5.1.12. *Management of products at the end of study*

At the end of the study and after the treatments are checked by the clinical research associate (CRA), the unused treatments may be destroyed on site, ONLY after written authorization from the sponsor (coordinating IHP).

##### 5.1.13. *In the event of batch recall*

If there is a batch recall for the product being studied, the sponsor will be informed by the Agence Nationale de Sécurité du Médicament et des produits de santé (French Agency for the Safety of Medicines and Health Products – ANSM). The sponsor will then inform the pharmacies at all research centers (by fax with proof of receipt and by letter) and will organize the quarantine, recall of the products, and their replenishment, if necessary.

#### 5.2. Associated Care and Treatments

Outside the specific treatments and investigations for the study, the treatment and follow-up are carried out as standard in compliance with clinical recommendations and standard procedures.

An ECG is performed after the second dose of the investigational drug. Analysis of this ECG by the physician in charge of the patient and, if needed, by a cardiologist, should be performed before the administration of the third dose of treatment:

- The QTc interval should remain ≤ 480 ms in the absence of continued ECG monitoring.
- If 480 ms < QTc < 500 ms, the patient must be monitored more closely (especially if bradycardia or monomorphic premature ventricular contractions occur regularly).
- If the QTc is ≥ 500 ms, this value must be confirmed by a new ECG, by using other correction formulas (e.g. Fridericia's formula), and, ideally, by an opinion from a specialist. Treatment should be reduced or stopped depending on the clinician's decision, and continuous cardiac monitoring implemented until ECG normalization.

Prescription of the lopinavir/ritonavir compound (in compliance with Decree n°2020-314 of March 25, 2020), azithromycin, and/or corticosteroids are authorized if the clinician deems it necessary. All instances of taking these medications during the period of participating in the study must be mentioned during the follow-up visits and will be taken into account in the analysis.

Prescribing azithromycin is authorized, if an antibiotic is prescribed, subject to following the Summary of Product Characteristics and the recommendations of the Réseau Français des Centres régionaux de Pharmacovigilance (French network of regional pharmacovigilance centers) (22 March 2020) (French only: https://www.rfcrpv.fr/chloroquine-point-dinformation).

In patients treated with azithromycin or concomitant medications prolonging QTc, or with drugs increasing hydroxychloroquine concentration, additional ECG should be carried out twice per week for the duration of treatment and in case of symptoms that could indicate problems with heart rate (sudden and short palpitations, fainting, ictal crisis, etc.). Action to be taken in the event of an increase in QTc will follow the same rules as those mentioned above.

With the exception of emergency situations that need specific treatments, the researcher must conduct the study in compliance with the protocols approved by the sponsor and the regulatory authorities.

In the event of complications that require intubation and invasive ventilation, the doctor responsible for the patient may request, and be granted, unblinding so that the patient can receive the most suitable treatments.

#### 6. Research Process

##### 6.1. Pre-selection/Recruitment

Participation in the study is offered to patients referred to or hospitalised in the participating centers.

##### 6.2. Inclusion Procedure

During the inclusion visit, if the patient meets the inclusion criteria for the study, written and verbal information is delivred by the investigator (information letter written in language that the patient can understand), who also answers any questions the patient may have. If the patient agrees to participate, the informed consent form is signed in three copies by all parties.

In the specific context of the COVID-19 pandemic, it is very likely that there will not be any relatives or legal representatives accompanying older patients who are suffering from COVID-19. Given this particular exceptional context and the need to administer the study drugs as soon as possible after diagnosis to limit the risk of worsening, the plan is to include patients who are not in a state to provide consent in the context of an emergency inclusion procedure (article L.1122-1-3 of the French Public Health Code (Code de Santé Publique). Inclusion (information and collecting consent) can therefore be carried out with a present representative of the patient. If there is no representative present, inclusion is carried out by the investigator as part of an emergency inclusion procedure (without consent). A representative of the patient is informed as soon as possible and he/she is asked for consent to continue with the study.

In both cases, the representative to request consent from is, in order of priority: legal guardian, legal supervisor, authorized person of trust, family member, or, otherwise, a person supporting the patient who has close and stable links.

In the event of inclusion as part of an emergency procedure, the patient is informed as soon as possible, if he/she regains his/her ability to provide consent, and he/she is asked for consent to continue with the study until the end of his/her participation in the study.

Inclusion in the context of an emergency procedure is indicated, if necessary, by specifying the means (inclusion by a representative of the patient or inclusion without consent, if no representative is present at the moment of inclusion). The dates of providing information and collecting signatures from the various parties (patient and patient representative if applicable, initial consent or consent to continue if applicable) are recorded in the patient's case file.

#### 6.3. Monitoring of Persons Participating in the Study

##### *6.3.1. Inclusion Visit*

The patient is randomized immediately after inclusion, then the investigational product is given to the patient, and administered in line with the defined protocol (cf. chapter 5).

The sociodemographic data, medical history, clinical examinations, and biological results are collected.

In the context of the ancillary study (200 patients), 37 ml of blood (one dry tube of 4 ml, four EDTA tubes, three of which contain 6 ml, three citrate tubes of 2.7 ml) are taken from patients who have agreed to take part in the biological collection (ancillary objective, cf. chapters 2 and 6.4) in the centers participating in this ancillary study. The tubes are disinfected and then placed in typical packaging before being taken to the laboratory via the usual channels (authorized pneumatic).

##### *6.3.2. ECG after the loading dose*

An ECG is performed after the second dose of investigational drug in order to check for rhythm abnormalities and QTc prolongation. Pursuit of the investigational drug is conditioned by the analysis of this ECG.

##### 6.3.3. Visit at day 3 ± 1

In patients taking concomitant medications prolonging QTc, or drugs increasing hydroxychloroquine concentration, an ECG is performed in order to check for rhythm abnormalities and QTc prolongation. In case of QTc prolongation, the investigator should follow the rules in paragraph 5.2.

##### 6.3.4. Visit at day 5 ± 1

***A nasopharyngeal swab sample*** is taken specifically for the needs of the study during a routine visit (cf. chapter 2.4.2), if and only if the patient was included on a positive RT-PCR result. If possible, the sample is analyzed in the center where the patient was included. If this is not possible, the sample is sent to the study sponsor's center (Angers University Hospital). This sample is taken only for patients whose COVID-19 diagnosis was confirmed by RT-PCR.

In patients taking concomitant medications prolonging QTc, or drugs increasing hydroxychloroquine concentration, an ECG is performed in order to check for rhythm abnormalities and QTc prolongation. In case of QTc prolongation, the investigator should follow the rules in paragraph 5.2.

In the context of the ancillary study, 37 ml of blood (one dry tube of 4 ml, three EDTA tubes of 6 ml, three citrate tubes of 2.7 ml and one EDTA tube of 6 ml) is taken from patients who have agreed to take part in the biological collection (ancillary objective, cf. chapters 2 and 6.4).

##### 6.3.5. Visit at day 7 ± 1

In patients taking concomitant medications prolonging QTc, or drugs increasing hydroxychloroquine concentration, an ECG is performed in order to check for rhythm abnormalities and QTc prolongation. In case of QTc prolongation, the investigator should follow the rules in paragraph 5.2.

##### 6.3.6. Visit at day 10 ± 1

***A nasopharyngeal swab sample*** is taken specifically for the needs of the study during a routine visit (cf. chapter 2.4.2), if and only if the patient was included on a positive RT-PCR result. If possible, the sample is analyzed in the center where the patient was included. If this is not possible, the sample is sent to the study sponsor's center (Angers University Hospital). This sample is taken only for patients whose COVID-19 diagnosis was confirmed by RT-PCR.

In patients taking concomitant medications prolonging QTc, or drugs increasing hydroxychloroquine concentration, an ECG is performed in order to check for rhythm abnormalities and QTc prolongation. In case of QTc prolongation, the investigator should follow the rules in paragraph 5.2.

In the context of the ancillary study, 37 ml of blood (one dry tube of 4 ml, three EDTA tubes of 6 ml, three citrate tubes of 2.7 ml and one EDTA tube of 6 ml) is taken from patients who have agreed to take part in the biological collection (ancillary objective, cf. chapters 2 and 6.4).

##### 6.3.7. Visit at day 14

A visit or telephone call is carried out to collect information on the occurrence of relevant clinical events at day 14 (cf. chapter 2). Information on the treatments received over the last 14 days are collected.

##### 6.3.8. *Visit at day 28*

A visit or telephone is carried out to collect information on the occurrence of relevant clinical events at day 28 (cf. chapter 2).

Information on the treatments received over the last 14 days are collected.

##### 6.3.9. *Special case of ambulatory patients*

For patients whose care is continued outside the center (at home or in another care facility), the following conditions must be met:

- The investigator must ensure prior to the exit that all the conditions for follow-up are met;
- The care environment and staff, including the attending physician, must be informed of the patient's participation in the study and the patient's modalities;
- Patient to attend scheduled follow-up visits;
- Systematic telephone follow-up is conducted by the center every 24-48 hours until J14. In case of doubt about the health of the person, an additional consultation at the hospital with an investigating doctor is proposed.

##### 6.3.10. *Summary table*

**Study Process**

| | Day 0 | Day 0-Day 1 | Day 3 ± 1 | Day 5 ± 1 | Day 7± 1 | Day 10± 1 | Day 14 | Day 28 |
|---|---|---|---|---|---|---|---|---|
| Consent/inclusion/randomization | X | | Finding consent to continue, if required | | | | | |
| Treatment administration | X | X | X | X | X | (until day 9) | | |
| Collecting sociodemographic, clinical, biological, and therapeutic data | X | | | X | | X | X | X |
| Electrocardiogram | X[1] | X | X[2] | X[2] | X[2] | X[2] | | |
| Kalaemia | X[1] | | | X[2] | | | | |
| Nasopharyngeal swab sample for RT-PCR | | | | X[3] | | X[3] | | |
| Biological samples for ancillary study* | X | | | X | | X | | |
| Observation and feedback on treatment | | | | | | X | | |
| Collecting adverse events and serious adverse events** | X | | X | X | X | X | X | X |
| Collecting patient status on WHO Ordinal Scale for Clinical Improvement ** | | | | | | | X | X |

[1]Prior to inclusion
[2]For patients receiving concomitent treatment interacting with hydroxychloroquine or prolonging QTc
[3]For patients with positive RT-PCR at inclusion
*for some centers
**consultation ou telephone follow-up if the patient has left the hospital.

#### 6.4. Collecting Biological Samples

For patients who agree to collection of biological samples, the samples taken for this purpose will be kept for 15 years, in an encoded and protected form, and held in the Biological Resource Center (BRC) at Angers University Hospital from the end of this study in the "Infectious Diseases" collection (DC-2016-2700, AC-2017-2993).

The sample tubes for biocollection are sent to the BRC at centers participating in the ancillary study, where they will be centrifuged and aliquoted according to the laboratory's practice. The aliquots of cells, plasma, and serum will be kept at -80°C, or at -196°C for mononuclear cells, in the local BRC and will then be sent to the BRC at Angers University Hospital at the end of the study.

The remaining samples from the ancillary study may be used or passed on for other research works (especially analyses of genetic characteristics on DNA and RNA) in order to improve scientific knowledge of issues linked to the COVID-19 infection.

Research works using the samples may be conducted by the team at Angers University Hospital only or a collaboration with other public, private, national, or international partners.

##### 6.5. Withdrawing a Person From the Study

Persons participating in the study can request to leave the study at any time, for any reason.

The researcher may either temporarily pause or permanently end a person's participation, or just their treatment as defined by the protocol, for any reason that would best serve the interests of that particular person in case of serious adverse events or a new factor arising in the study. In the event that the treatment as defined by the protocol is stopped, the researcher must ensure an adequate treatment and, to the extent possible, the continued follow-up as planned in the study.

In the event of loss of contact, the investigator will do everything possible to regain contact with the person and to ensure the planned continuation of the study. In case of early withdrawal, the investigator will document the reasons for this in as much detail as possible.

People with whom contact is lost or who withdraw early from the study will not be replaced.

The data captured for people lost to follow-up or who withdraw early from the study will be used in the analysis stage, including in cases where consent is withdrawn (unless the person exercises their right to have the personal data erased). If the patient demands that their data be erased, the data required to evaluate the safety data will be kept.

##### 6.6. Research Duration

Inclusion period duration: 4 months maximum

Study participation duration for a participant: 28 days

Total study duration (inclusion duration + participation duration): 5 months maximum

#### 7. Risk-Benefit Balance

The predicted risks and constraints for participants, as described in the paragraphs below, are acceptable in light of the expected benefits.

##### 7.1. Benefits

###### *7.1.1. Individual benefit*

For the study participants taking Plaquenil®, the anticipated effect is a reduction in the risk of complications and in the use of intubation and artificial ventilation and/or death.

##### *7.1.2. Collective benefit*

The anticipated benefit is considerable as it answers the question of Plaquenil®'s efficacy in preventing complications in patients suffering from COVID-19 who are at risk of their condition worsening. There is currently no controlled, randomized study that can confirm the efficacy of Plaquenil® in this indication. However, this drug leads to some adverse effects and it is important to confirm its real efficacy before rolling out its use in routine treatment for COVID-19 patients. From the point of view of public health, there is serious interest in identifying an effective treatment during this COVID-19 pandemic.

#### 7.2. Risks

Taking the nasopharyngeal swab samples may cause some pain.

In the placebo group, treatment is identical to that of patients who are not participating in this study. The risk faced by patients participating in the study is not benefiting from a treatment that may prove to be effective.

In the Plaquenil group, the risks are linked to the side effects of the treatment. These are rare overall if the treatment is for a short term. The main contraindications are checked before patients are asked to participate in this study (known intolerance, combination of certain drugs). An electrocardiogram (ECG) is carried out on inclusion to ensure that there is no QT interval prolongation in any of the patients evaluated in preparation for taking part in the study. Another ECG is performed after the loading dose in order to ensure that the drug does not prolong QTc. In patients at higher risk of cardiac toxicity because of concomitant drug interacting with hydroxychloroquine or prolonging QTc, additional ECG are performed every 48 to 72 hours. Common side effects are listed for patients in the information letter and they are asked to inform their doctor of any unusual symptoms.

### 8. Safety Assessment

All adverse events, both serious and minor, arising during this study will be dealt with in compliance with the procedures established by Angers University Hospital, in accordance with current regulations (cf. appendix 1).

#### 8.1. Description of the Safety Assessment Parameters

The safety of patients is assessed specifically in the context of this study through monitoring:

- clinical factors: electrocardiogram prior to inclusion in all patients and every 48 hours for at-risk patients (treatment that could lengthen QTc interval), monitoring and collecting information on adverse events during each visit

- biological factors: potassium levels in patients at risk of hypokalemia due to medication, blood sugar levels in the event of indicative clinical signs.

Monitoring kidney function is done in a non-specific manner.

Information on treatments that could interact with hydroxychloroquine will be collected.

#### 8.2. Declaration of Adverse Events: Protocol Specificity

- All adverse events that occur during a patient's participation will be declared to the sponsor, excluding adverse events linked to the COVID-19 infection.

Serious adverse events must be declared to the sponsor immediately and no later than 24 hours from the moment when it becomes known (cf. appendix 1).

The following adverse events, which are mentioned in the Summary of Product Characteristics (SmPC) and are potentially linked to the investigational drug, will be specifically assessed:

- Abnormal heart rhythm, cardiac conduction disorders
- Seizures
- Visual disturbances
- Hypoglycemia
- Skin rash or pruritus
- Recurrent vomiting

Every other event potentially linked to the treatment will be recorded.

The events linked to COVID-19 infection will not be recorded as an adverse event in the CRF, nor declared as a serious adverse event to the Pharmacovigilance Officer.

In addition, the following conditions linked to COVID-19 infection will not be declared to the Pharmacovigilance Officer:

- Rhinitis
- Anosmia/dysgeusia/ageusia
- Fever
- Cough
- Confusion
- Lymphocytopenia
- Acute respiratory distress syndrome
- Multiple organ dysfunction
- Death
- Hospitalization or extended hospitalization

* Given the short duration of treatment, the likelihood of a pregnancy occurring during the study is very low. If necessary, an adverse event will be declared with the ad hoc form. The study treatment will be stopped. Unblinding may take place at the request of the doctor who is responsible for the patient. Pregnancies will be monitored by the sponsor to full term.
* Serious adverse effects that are life-threatening or fatal will be sent by the sponsor to ANSM, as will suspected unexpected serious adverse reactions (SUSARS – serious abnormal heart rhythm, heart block disorders, or other heart conditions).

#### 8.3. Reference Documents

The reference document used to evaluate the expected/unexpected character of adverse events is the Summary of Product Characteristics for Plaquenil® (hydroxychloroquine). The Reference Safety Information (RSI) corresponds to chapter 4.8 of the Summary of Product Characteristics.

As the information given in the marketing authorizations is subject to change, it should be ensured at the time of prescribing the drug to respect, in particular, contraindications, warnings and precautions for use, drug interactions, and methods of contraception. Refer to the information available on the French Public Drug Database, which can be accessed online at the following website (French only): http://base-donnees-publique.medicaments.gouv.fr/
A list of drugs interacting with hydroxychloroquine is available in the *Thésaurus des Interactions Médicamenteuses* from the *Agence Nationale de la Sécurité du Médicament* (https://www.ansm.sante.fr/Dossiers/Interactions-medicamenteuses/Interactions-medicamenteuses/(offset)/0).

##### 8.4. Independent Data and Safety Monitoring Board

An independent monitoring board (IDSMB) has been put together for this study. A charter defines the primary responsibilities of the independent monitoring committee, its composition, its objectives, relationships with other parties in the study, and the frequency of its meetings.

#### 9. Statistics

Quantitative variables will be recorded as an average +/- standard deviation or median and interquartile range for non-normal distributions. They will be recorded as both numbers and percentages. Averages will be compared using the Student's t-test (or the Mann-Whitney test if required) and percentages will be compared using the Chi-squared text (Fisher's exact test if required).

The main analysis will be performed in the intention-to-treat population. The null hypothesis of the study is that there is no difference between the rate of the composite of use of intubation and mechanical ventilation or death at day 14 between hydroxychloroquine and placebo.

Due to the current exceptional circumstances, interim analyses will be performed every 50 patients to compare the two groups based solely on the main evaluation criterion. The frequency of interim analyses will be adapted depending on the number of inclusions and gathering of analyzable findings based on the main evaluation criterion but will not be changed more than once a day. At the end of each meeting, the IDSMB will determine the date for the interim analysis to be performed and for the subsequent meeting. A unilateral alpha risk will be maintained at 2.5% during these interim analyses using the unilateral triangle test. If the upper limit of the triangle is exceeded, the test will conclude to the efficacy of hydroxychloroquine. If the lower limit of the triangle is exceeded, the test will conclude to the futility of hydroxychloroquine (54).

The secondary study criteria will only be analyzed at the end of the study:

- the mortality rate or use of invasive ventilation in the 28 days (day 28) following inclusion and the start of treatment will be compared in both groups using the Chi-squared test (or Fisher's exact test if required)

- the clinical evolution via the WHO Ordinal Scale for Clinical Improvement at day 14 and day 28 will be compared in both groups using the Mann-Whitney test

- the all-cause mortality at day 14 and day 28 will be compared in both groups using the Chi-squared test (or Fisher's exact test if required); survival curves will be established between day 0 and day 28 using the Kaplan-Meier method

- the virus transmission assessed by the rate of RT-PCR positive for SARS-CoV-2 with respiratory tract samples at day 5 and day 10 will be compared in both groups using the Chi-squared test (or Fisher's exact test if required).

- the incidence of symptomatic venous thrombo-embolism occurrences at day 14 and day 28 will be compared in both groups using the Chi-squared test (or Fisher's exact test if required).

- the rate of serious adverse events at day 28, defined in accordance with current regulations, will be analyzed using the Chi-squared test (or Fisher's exact test if required) in the safety population of patients having received at least one dose of the allocated treatment.

- for the ancillary study, biological markers will be recorded as an average +/- standard deviation or median and interquartile range for non-normal distributions. Averages will be compared using the Student’s t-test and medians will be compared using the Mann-Whitney test.

A sub-group study will be carried out among people aged 75 years or older. This will focus on the clinical development using the OSCI scale and on all-cause mortality at day 14 and day 28.

The significance threshold will be set at 0.05. Sequential analysis based on the triangle test will be performed using R version 3.6.3 software and Stata software (version 13) will be used for other analyses.

### 10. Data Management

#### 10.1. Data Collection Methods

All data related to this study are collected using a standardized electronic case report form (eCRF) and on the basis of valid documents (patient medical record). In the eCRF, follow-up calls, and monitoring forms, patients can be identified by a unique number composed of the center number and the patient number at the center. The initials of the patient’s name and surname are also gathered.

Due to the current exceptional circumstances, it is expected that some centers will be unable to provide a follow-up call at day 14 and day 28. For centers unable to provide telephone follow-up for patients included on day 14 and day 28, telephone follow-up will be provided by Angers University Hospital. Angers University Hospital obtained an authorization (no. 920125) from the French Data Protection Authority (CNIL, *Commission nationale de l'informatique et des libertés*) on april 3rd 2020.

For these centers unable to provide the follow-up, the personal data needed to perform the centralized monitoring is collected on a paper form and faxed to the centralized monitoring team. The data collected during centralized monitoring is entered into the eCRF. The original copies of the centralized follow-up questionnaires are sent by the coordinating center to the investigation centers regularly throughout the study in order to store patient medical records.

The confidentiality of patients and their personal health information is maintained by restricting access to patient records and eCRF at all times. Identifiable data shall only be accessible to the study researchers and authorized personnel responsible for the running of the study, as well as to evaluators assigned by the sponsor to ensure the accuracy of the data, and to other members of regulatory bodies that are legally authorized to have access to the data. Lastly, any disclosure of confidential data required by applicable laws and regulations is carried out where necessary.

The eCRF (Ennov Clinical) is managed by the Delegation for Clinical Research and Innovation (DRCI) of Angers University Hospital. It is accessible via web browser.

##### 10.2. Data Collection and Confidentiality

Individuals who have direct access to the data will take all necessary precautions to ensure the confidentiality of information relating to the data of trial participants, particularly in relation to their identity and the results obtained. Those with access are subject to professional confidentiality (according to the terms of articles 226-13 and 226-14 of the French Penal Code).

The data is encrypted and stored. Study participants can be identified by a number composed of the center number and a sequential inclusion number given by the center. Only the first letters of the patient's name and surname will be recorded, along with the month and year of their birth.

A mailing list is kept by each center and falls under the responsibility of the principal investigator of the center. This list is kept for the prescribed duration for this type of study.

##### 10.3. Right to Access Data and Source Documents

In accordance with Good Clinical Practices:

- the sponsor ensures that each person participating in the study has given their written consent for access to their personal data that is strictly necessary for the quality control of the study.

- the researchers make the documents and individual data strictly necessary for this control available to the persons responsible for monitoring, quality control, auditing of the research or, where appropriate, inspections by the relevant authorities.

##### 10.4. Quality Control and Assurance

A level of monitoring is determined according to the risks of the study, as defined by the sponsor's procedures. The monitoring guide details the follow-up methods defined by the sponsor.

Due to the situation at the time of study start with prolonged lockdown in France, on-site monitoring is not feasible. For all that, continuous monitoring is essential as interim analyses are performed every 50 patients. Hence, the sponsor will organize remote consent inspections by clinical research associates as long as needed. The secure process implemented ensures that consents sent by the centers are accessible only by the CRAs, who are the only representatives of the promoter authorized to know the identity of the participants.

After verification, the consents are placed in sealed envelopes with the signed list and kept locked up in the CRA office until the study is archived.

In the same way, remote monitoring is set up to control the data of the main judgment criterion on D14 and to ensure the safety of patients on the basis of hospitalization and consultation reports.

The documents recovered during the remote monitoring will be destroyed at the end of the study.

On-site monitoring will complement remote monitoring at the end of the health emergency period.

The monitoring methods proposed for this study come from the recommendations of the European drug agency "Guidance on the management of clinical trials during the COVID-19 pandemic (V2 of March 27, 2020).

##### 10.5. Archiving

The researcher and the sponsor ensures the preservation of documents and data related to the study in accordance with current regulations. The methods used to preserve these essential documents ensure that the documents remain complete and legible for the entire period for which their preservation is required.

The investigators are responsible for the safe keeping of documents essential to the study at the research site. If they leave the institution, they must inform the sponsor and delegate the remainder of the safe keeping in writing.

#### 11. Ethical and Regulatory Considerations

##### 11.1. Study Classification

This is an interventional drug study.

##### 11.2. Ethics Committee and Relevant Authorities

The study file (notably the protocol, the abstract, the information sheet, and the consent form) have been submitted to the *Comité de Protection des Personnes du Sud-Ouest et Outre-Mer 4* (CPP SOOM4), which approved this study on March 30, 2020 (N° CPP SOOM4 2020-03-036 / N° SI CNRIPH 20.03.24.72431). The *Agence Nationale de Sécurité du Médicament et des Produits de Santé* (ANSM) authorized this research on March 31, 2020 (N° MEDAECNAT-2020-03-00045).

In cases where substantial changes are to be made to the study file by the researcher, the sponsor will seek approval from the CPP and authorization from the ANSM before making such changes. Renewed consent from persons participating in the study will be obtained if required.

The study end date will be sent by the sponsor to ANSM and the IRB within 90 days. The research end date corresponds to the end of participation of the last study participant, or, if applicable, to the theoretical end date defined by the protocol (if the prescribed objective is not achieved and without request for extension).

##### 11.3. Processing of Personal Data

Data management in this study falls within the framework of "Reference Methodology" (MR-001) in accordance with the provisions of the amended law no. 78-17 of January 6, 1978 in relation to data processing, files and civil liberties (French Data Protection Act). Angers University Hospital, the study sponsor, has signed an agreement to comply with this "Reference Methodology" (declaration number 1174822).

The data management of this study is performed in accordance with the European General Data Protection Regulation, which came into effect on May 25, 2018, and with the French Data Protection Act.

An authorization was obtained on april, 3rd 2020 from the French Data Protection Authority (CNIL) to guarantee the centralized follow-up of patients from centers that cannot provide this.

The amended protocol describing the specific monitoring procedures during the health crisis will also be submitted to the CNIL.

##### 11.4. Protocol Specifics

###### *11.4.1. Simultaneous participation in another study and exclusion period definition*

Simultaneous participation in another interventional study that changes methods of treatment is not allowed during the entire participation period of the study (28 days). The protocol does not indicate an exclusion period at the end of participation during which individuals must not participate in another interventional study.

###### *11.4.2. Indemnity of people participating in the study*

No indemnity as compensation for stress incurred is foreseen for study participants.

###### *11.4.3. Insurance*

The sponsor has taken out an insurance policy for the entire duration of the study, guaranteeing its own civil liability and that of all doctors and collaborators involved in the study (insurance provided by SHAM, *Société Hospitalière d'Assurances Mutuelles*: policy number 147412). It shall also provide full compensation for any harmful consequences of the research for the participants or their beneficiaries, unless it is able to prove that the damage is not attributable to its fault or to that of any person involved in the trial, without the possibility of holding against them a third party's actions, or voluntary withdrawal from the trial by a person who had initially consented to take part in the research.

#### 12. Rules Relating to Publication

The communications and scientific reports corresponding to this study fall under the responsibility of the coordinating researcher for this study, in collaboration with the chief co-researchers and associated scientists. The co-authors of the report and publications are the researchers and the clinicians involved, in proportion to their contribution to the study, as well as the methodologist and/or the biostatistician and associated researchers.

The first author will be principal researcher V. Dubée, the second author P.-M. Roy, the penultimate author I. Pellier, and the last author A. Mercat.

The primary results of the study will be subject to a final report and publication and/or specifically related scientific presentation. Except with specific authorization, no phase of the study may be the subject of an oral presentation or poster before acceptance of the corresponding manuscript in a referenced journal.

Angers University Hospital should be referenced in all publications regarding this study.

The study was registered on an open-access website (*ClinicalTrials.gov*) before the inclusion of the first patient in this study (NCT04325893).

#### 13. Références

1. Zhu N, Zhang D, Wang W, Li X, Yang B, Song J, et al. A Novel Coronavirus from Patients with Pneumonia in China, 2019. N Engl J Med. 2020 20;382(8):727–33.

2. Qin C, Zhou L, Hu Z, Zhang S, Yang S, Tao Y, et al. Dysregulation of immune response in patients with COVID-19 in Wuhan, China. Clin Infect Dis Off Publ Infect Dis Soc Am. 2020 Mar 12;

3. Chen J, Wang X, Zhang S, Liu B, Wu X, Wang Y, et al. Findings of Acute Pulmonary Embolism in COVID-19 Patients [Internet]. Rochester, NY: Social Science Research Network; 2020 Mar [cited 2020 Mar 23]. Report No.: ID 3548771. Available from: https://papers.ssrn.com/abstract=3548771

4. Zhou F, Yu T, Du R, Fan G, Liu Y, Liu Z, et al. Clinical course and risk factors for mortality of adult inpatients with COVID-19 in Wuhan, China: a retrospective cohort study. Lancet Lond Engl. 2020 Mar 11;

5. Zuin M, Rigatelli G, Zuliani G, Rigatelli A, Mazza A, Roncon L. Arterial hypertension and risk of death in patients with COVID-19 infection: systematic review and meta-analysis. J Infect. 2020 Apr 10;

13. Kravvariti E, Koutsogianni A, Samoli E, Sfikakis PP, Tektonidou MG. The effect of hydroxychloroquine on thrombosis prevention and antiphospholipid antibody levels in primary antiphospholipid syndrome: A pilot open label randomized prospective study. Autoimmun Rev. 2020 Feb 19;102491.

31. multicenter collaboration group of Department of Science and Technology of Guangdong Province and Health Commission of Guangdong Province for chloroquine in the treatment of novel coronavirus pneumonia. [Expert consensus on chloroquine phosphate

for the treatment of novel coronavirus pneumonia]. Zhonghua Jie He He Hu Xi Za Zhi Zhonghua Jiehe He Huxi Zazhi Chin J Tuberc Respir Dis. 2020 Feb 20;43(0):E019.

32. Cortegiani A, Ingoglia G, Ippolito M, Giarratano A, Einav S. A systematic review on the efficacy and safety of chloroquine for the treatment of COVID-19. J Crit Care. 2020 Mar 10;

46. Nomura A, Kishimoto M, Takahashi O, Deshpande GA, Yamaguchi K, Okada M. Prolongation of heart rate-corrected QT interval is a predictor of cardiac autonomic

dysfunction in patients with systemic lupus erythematosus. Rheumatol Int. 2014 May;34(5):643–7.

47. Costedoat-Chalumeau N, Amoura Z, Duhaut P, Huong DLT, Sebbough D, Wechsler B, et al. Safety of hydroxychloroquine in pregnant patients with connective tissue diseases: a study of one hundred thirty-three cases compared with a control group. Arthritis Rheum. 2003 Nov;48(11):3207–11.

48. 050710s039,050711s036,050784s023lbl.pdf [Internet]. [cited 2020 Mar 19]. Available from: https://www.accessdata.fda.gov/drugsatfda_docs/label/2013/050710s039,050711s036,050784s023lbl.pdf

49. Centre de Référence des Agents Tératogènes. Hydroxychloroquine - Grossesse et allaitement [Internet]. Available from: https://lecrat.fr/articleSearchSaisie.php?recherche=plaquenil

50. Petri M. Pregnancy and Systemic Lupus Erythematosus. Best Pract Res Clin Obstet Gynaecol. 2019 Oct 8;

51. Mekinian A, Alijotas-Reig J, Carrat F, Costedoat-Chalumeau N, Ruffatti A, Lazzaroni MG, et al. Refractory obstetrical antiphospholipid syndrome: Features, treatment and outcome in a European multicenter retrospective study. Autoimmun Rev. 2017 Jul;16(7):730–4.

52. Yao X, Ye F, Zhang M, Cui C, Huang B, Niu P, et al. In Vitro Antiviral Activity and Projection of Optimized Dosing Design of Hydroxychloroquine for the Treatment of Severe Acute Respiratory Syndrome Coronavirus 2 (SARS-CoV-2). Clin Infect Dis. 2020 Mar 9.

53. WHO. R&D Blueprint - novel Coronavirus - COVID-19 Therapeutic Trial Synopsis [Internet]. WHO, Geneva; 2020 [cited 2020 Mar 23]. Available from: https://www.who.int/blueprint/priority-diseases/key-action/COVID-19_Treatment_Trial_Design_Master_Protocol_synopsis_Final_18022020.pdf

54. Whitehead J. The Design and Analysis of Sequential Clinical Trials, Revised, 2nd Edition | Ed. Ellis Horwood, (1992)

#### 14. Appendices

Appendix 1: Safety Appendix

Appendix 2: Methods of administration of the study treatment by enteral feeding tube

### Appendix 1: Safety of the Clinical Trial

**1 - Definitions**

- **Adverse event (AEv):** any untoward medical occurrence that affects a person taking part in an interventional study, whether or not this event is linked to the study or to the investigational product(s) that this study concerns.
- **Serious adverse event (SAEv):** the severity is defined by one of the following observations:
  - Death
  - Endangering life (immediately life threatening at the moment of the event, and this, independent of the consequences that would have a corrective or palliative therapeutic impact)
  - Significant or sustained incapacity or disability
  - Hospitalization
  - Extension of hospitalization
  - Congenital malformation/anomaly
  - Potentially serious event (adverse clinical event or laboratory result of a serious nature or considered as such by the researcher)
- **Adverse effect (AE):** any harmful and adverse reaction to an investigational product, regardless of the dose administered.
- **Serious adverse effect (SAE):** serious adverse event due to an investigational product.
- **Unexpected adverse effect (UAE):** adverse effect whose nature, severity, intensity or evolution is not consistent with the information in the summary of characteristics for an authorized investigational product or, in the case of a non-authorized drug, in the brochure of the investigator.
- **Imputability:** relation between the AEv and the treatment in the study. The AEv linked to an investigational product shall become an AE. The factors to take into account for determining imputability are:
  - the chronology of events,
  - the disappearance of the AEv on stopping the drug and/or its reappearance in the case of re-administration,
  - the pharmacodynamics and pharmacokinetics of the drugs,
  - the notion of a similar event occurring previously on administration of the drug or of a drug of the same class
  - the existence of another etiology.

- **Intensity**: the intensity of the AEv is evaluated by the researcher, either through the use of a rating scale for adverse events attached to the protocol (e.g: NCI-CTC classification for oncology trials), or according to the following classification:
    - mild grade 1: AEv generally transitory and without impact on normal activities
    - moderate grade 2: AEv sufficiently compromising to have an effect on normal activities
    - severe grade 3: AEv considerably changing, or impeding, the person's normal course of activities or constituting a danger to the life of the person

  N.B.: the intensity criteria should not be confused with the severity criteria that serves as a guide for defining the reporting obligations.

#### 2 - Role of the Researcher

##### 2.1 - Notification of Adverse Events

###### *2.1.1 - Information to relay to the sponsor*

Each SAEv shall be described on the form to this effect ("Declaration of Serious Adverse Event - Drug") and endeavor to provide as much detail as possible.

The researcher should also attach to the SAEv report, where possible:

- a copy of the hospital report or of the extended hospitalization notice,
- if relevant, a copy of the autopsy report,
- a copy of all the additional exam results carried out, including the relevant negative results and attaching the normal laboratory values,
- any other document considered useful or relevant.

These documents will be anonymized and marked with the identification number of the patient.

Each adverse event will be monitored until it is completely resolved (stabilization at a level judged to be acceptable by the researcher or return to the previous state), even if the patient has left the trial.

Certain circumstances that require a period of hospitalization do not fall under the "hospitalization / prolonged hospitalization" severity criteria and do not need to be classified as SAEv:

- hospitalization predetermined by the protocol
- admission for social or administrative reasons
- transfer to day clinic
- hospitalization for routine treatments or check-ups related to the studied condition, not caused by a deterioration in the patient's health
- hospitalization for medical or surgical treatment scheduled before the beginning of the trial

For non-serious adverse events, the "AE" page in the CRF is completed with the description of the event, the start and end dates of the event, the development, the severity, the link to study treatment, the intensity, measure(s) taken.

###### *2.1.2 - Methods for notifying the sponsor*

Every SAEv, whatever the causal relationship with the treatment(s) of the trial or the study (with the exception of those identified in the protocol as not needing an immediate declaration), must be sent by fax to +33 (0)2 41 35 59 68 or by e-mail to.

Non-serious adverse events are recorded by the researcher via the CRF.

###### *2.1.3 - Sponsor declaration period*

Without exception, every SAEv must be declared without delay (or as soon as the study doctor is made aware of it) to the study sponsor. Any additional information shall be sent by the researcher to the sponsor as soon as possible.

###### *2.1.4 - Sponsor notification period*

Excluding the specificities defined in paragraph 8.2. regarding the notification period, the researcher is responsible for recording and reporting every SAEv that occurs throughout the trial:

- from the date of signing the consent,
- throughout the trial's scheduled follow-up period of the participant

Furthermore, regardless of how long after the end of the trial it occurs, any SAEv potentially caused by the study must be declared to the sponsor, so long as there is no other cause that the study can reasonably attribute it to (for example, serious effects, such as cancers or congenital anomalies, can manifest long after exposure to the drug).

##### 2.2 - Patient Care

In the event of occurrence of a serious adverse event, the patient shall be treated according to the nature of the event and according to the habitual practices of the study center.

In the context of a blind trial, it can be unblinded if the medical care to be given is different depending on treatment administered.

In case of an adverse event having led to a patient leaving the trial or of an event persisting at the end of the trial, the patient shall be monitored until the event is resolved.

#### 3 - Role of Sponsor

##### 3.1 - Analysis of Serious Adverse Events

The sponsor must evaluate:

- the causality of the SAEv: all adverse events, for which the researcher or the sponsor believes that a causal relationship with the investigational product(s) can be reasonably assumed, are considered as suspicions of adverse effects. In cases where the evaluations by the sponsor and the study doctor differ, the two opinions are recorded in the declaration addressed to the relevant authority if this is deemed necessary,
- their expected or unexpected nature, taking recourse to the reference document in the protocol in force.

##### 3.2 - Imputability Score

In accordance with the ICH recommendations on the management of adverse events in clinical trials, a causality assessment is carried out for every SAEv declared. The rating method used is the following:

- **No relation**: the event appears within a period that does not fit in with the administration of the drug, and/or a sufficient amount of information exists to show that the reaction observed is unrelated to the drug, and/or there exists a plausible alternative explanation.
- **Doubtful relation**: the event has a chronology (appearance, development) that is slightly compatible with the administration of the drug and is plausibly attributable to factors other than the drug, such as the clinical state of the person or concomitant administration of other drugs.
- **Possible relation**: the event appears within a compatible period after administration of the drug, and although it is not possible to exclude the drug as a contributing factor, other factors can be brought into question, such as the clinical state of the person or the concomitant administration of other drugs. The information on the development on stopping the drug (dechallenge) can be absent or non-conclusive.
- **Probable relation**: the event appears within a compatible period after the administration of the drug. It cannot be reasonably attributed to another factor, such as the clinical state of the person or the concomitant administration of other drugs. The development on stopping the drug (dechallenge) must be clinically compatible. Information on the re-introduction of the drug (rechallenge) is not required.
- **Highly probable relation:** the event appears within a very suggestive period after the administration of the drug. It cannot be explained by another factor, such as the clinical state of the person or the concomitant administration of other drugs. The development on stopping the drug (dechallenge) must be clinically compatible. The event is explicable on a pharmacological or physiopathological level, or has recurred on the re-administration of the drug.

Adverse events with a doubtful, possible, probable, or highly probable relation with the investigational product(s) are considered to be linked. If they are unexpected, they qualify as being USAE and must be declared by the sponsor (see following paragraph).

##### 3.3 - Declaration of Unexpected Serious Adverse Effects

The sponsor declares all serious and unexpected serious adverse effects (USAE) to Eudravigilance (database of European pharmacovigilance), the French Agency for the Safety of Medicines and Health Products (ANSM), and to the researchers. The regulatory declaration should be made:

- **immediately** for unexpected serious adverse effects that are fatal or life threatening. In this case, the relevant complementary information must be gathered and sent in within a further **8 days**.

- **within 15 calendar days** for all other serious unexpected effects. Similarly, relevant complementary information must be gathered and sent within a further **8 days**.

In the case of a blind trial, as a general rule, the sponsor declares the unexpected serious adverse effect to the health authorities after having unblinded the investigational product.

In exceptional cases, and with the agreement of the ANSM solicited by the sponsor when applying for authorization of the clinical trial, the methods for unblinding and declaration of suspicions of adverse effects can be adjusted. *These methods are defined in detail in the research protocol (or in an attached document).*

The ANSM may request that the detailed records of all the adverse events recorded by the researchers be sent to them.

##### 3.4 - Transmission of Annual Safety Reports

On the anniversary date of the authorization of the trial issued by the health authorities, the sponsor drafts a safety report that includes:

- the list of serious adverse effects that are likely to be linked to the investigational product(s) in the trial, including the unexpected and expected serious effects.

- a concise and critical analysis of the safety of the patients involved in the study.

This report can be submitted to the coordinating researcher for approval. It must be sent to the relevant authorities (ANSM) and to the IRB within **60 days** of the anniversary date of the authorization of the trial.

##### 3.6 - Declaration of Other Safety Information

This involves all safety information or any new facts that could significantly modify the evaluation of the risk-benefit ratio of an investigational product or of the trial or that could lead to considering modifications concerning the administration of the drug or the carrying out of the trial, such as:

a) any significant clinical increase in the frequency with which an unexpected serious adverse effect occurs.

b) suspicions of USAE that occurred in participants who finished the trial and who are flagged to the sponsor by the researcher, as well as any follow-up reports.

c) any new fact concerning the course of the clinical trial or the development of the IP, when this new fact may put the safety of participants at risk. For example:

- a serious adverse event that is likely linked to investigations and to diagnostic procedures of the trial and could modify the course of this trial,

- a significant risk for the study population, such as a lack of efficacy of the IP used in the treatment of a life-threatening illness,
- significant and reliable results coming from a recently terminated study in animals (such as a study of carcinogenicity),
- an anticipated termination or temporary interruption of a trial being conducted with the same investigational product in another country for safety reasons
- a USAE linked to a non-experimental drug necessary for the performance of the trial (e.g.: challenge agents, fallback treatment)

d) recommendations of the independent monitoring committee, where appropriate, if they are necessary for the safety of those involved

e) any USAE sent to the sponsor by another sponsor of a clinical trial led in a third-party country that concerns the same drug.

The sponsor must declare this information to the ANSM and the relevant IRB as soon as possible. Additional relevant information must be sent within a further **8 days.**

### Appendix 2. Methods of administration of the study treatment by enteral feeding tube

- Put on medical gloves and a surgical mask
- Stop enteral nutrition and rinse the tubing with 20 – 30 ml of water
- Crush the study treatment in a clean mortar and resuspend in 10 – 15 ml of water
- Draw up into a large-tip syringe with a large tip and administer as a bolus, without any other associated product
- Rinse the tubing with 10 – 15 ml of water before potential administration of any other drug
- After administration of the last medication: rinse the tubing with 20 – 30 ml of water
- Resume feeding if necessary
