## Supplementary data for "A placebo-controlled double blind trial of hydroxychloroquine in mild-to-moderate COVID-19"

### Supplementary Figures

#### Figure S1. Patient selection, treatment allocation, and follow-up.

Assessment of eligibility criteria was performed in 33 of the 51 participating centers, and 202 patients were included in these centers (11%).

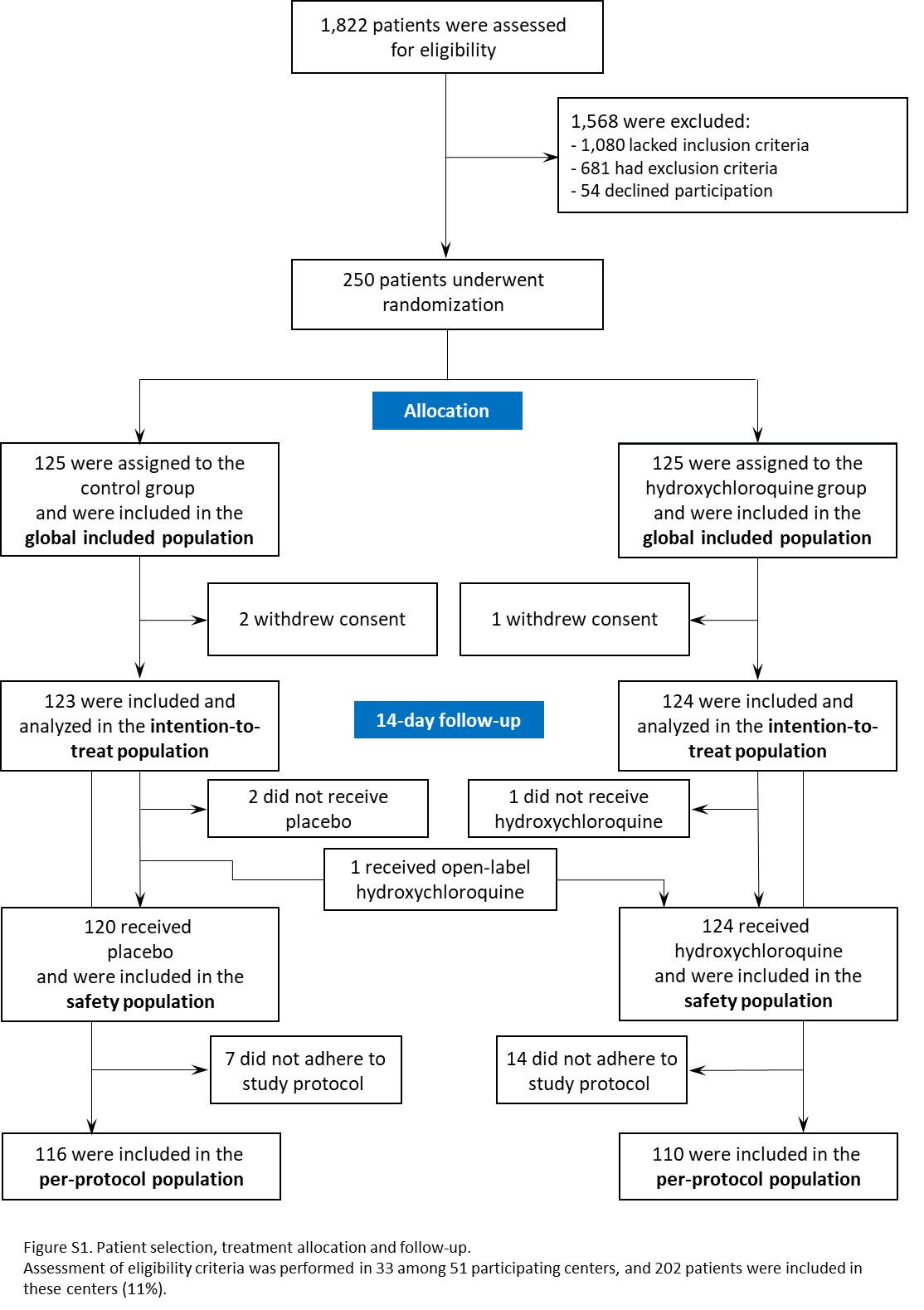

#### Figure S2. Age distribution of patients included in the study.

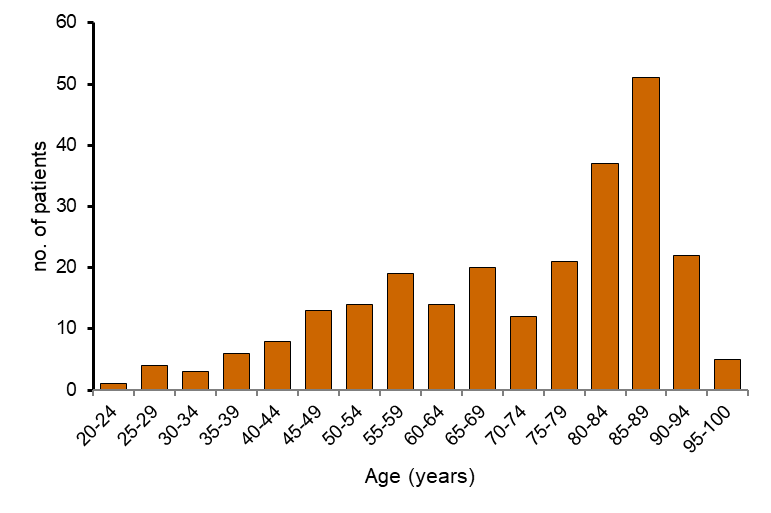

#### Figure S3. Kaplan–Meier estimation of primary endpoint-free survival.

HCQ, hydroxychloroquine.

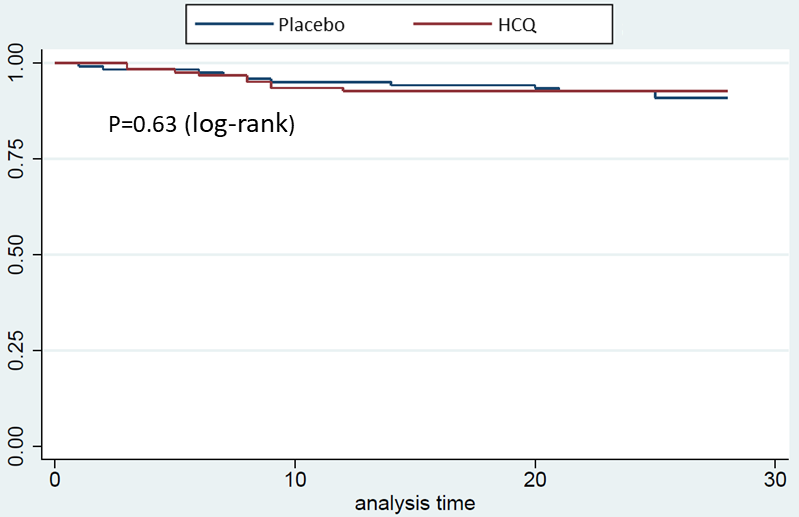

| Table S1. Major violations to the protocol leading to the exclusion of the per-protocol population. | | |
| --- | --- | --- |
| Major violations | | Number of patients* |
| Inclusion and exclusion criteria | |  |
|  | One or more inclusion criteria lacking | 3 |
|  | Presence of at least one exclusion criteria | 4 |
| Deviation in treatment administration protocol | |  |
|  | Allocated treatment not started | 3 |
|  | Allocated treatment prematurely stopped for reasons other than PE met or occurrence of serious AE, with less than 80% of the planned doses administered, or missing data on adherence to allocated treatment | 5 |
|  | Administration of hydroxychloroquine to a patient in the placebo group | 1 |
|  | Loading dose not correctly administered | 4 |
|  | Non-respect of the treatment regimen with a daily dose of the allocated treatment higher than expected (total number of tablets taken within less than 8 days) | 4 |
| PE, primary endpoint; AE, adverse event | |  |
| *In three patients, two deviations were observed. | |  |

### Supplementary Tables

| Table S2: Outcomes in the per-protocol population | | | | | | |
| --- | --- | --- | --- | --- | --- | --- |
|  |  |  | **All patients** | **Hydroxychloroquine** | **Placebo** | **Relative risk (95% CI)** |
| **Characteristics** | | | **(N = 247)** | **(N = 110)** | **(N = 116)** |  |
| Primary outcome | | |  |  |  |  |
|  | Day 14 | | 13 (5.8) | 7 (6.4) | 6 (5.2) | 1.23 (0.43–3.55) |
|  | Day 28 | | 17 (7.5) | 7 (6.4) | 10 (8.6) | 0.74 (0.29–1.87) |
| Mortality | |  |  |  |  |  |
|  | Day 14 | | 9 (4.0) | 5 (4.6) | 4 (3.5) | 1.32 (0.36–4.78) |
|  | Day 28 | | 14 (6.2) | 5 (4.6) | 9 (7.8) | 0.59 (0.20–1.69) |
| Use of intubation and mechanical ventilation | | | |  |  |  |
|  | Day 14 | | 4 (1.8) | 2 (1.8) | 2 (1.7) | 1.05 (0.15–7.36) |
|  | Day 28 | | 5 (2.2) | 2 (1.8) | 3 (2.6) | 0.70 (0.12–4.13) |
| Clinical evolution, as compared to day 0 | | | |  |  |  |
|  | Day 14 | |  |  |  |  |
|  |  | Absence of deterioration | 206 (91.2) | 100 (90.9) | 106 (91.4) | 1.00 (0.93–1.08) |
|  |  | Clinical improvement | 149 (65.9) | 73 (66.4) | 76 (65.5) | 0.97 (0.82–1.15) |
|  |  | Recovery | 127 (56.2) | 62 (56.4) | 65 (56.0) | 0.97 (0.78–1.22) |
|  | Day 28 | |  |  |  |  |
|  |  | Absence of deterioration | 210 (92.9) | 103 (93.6) | 107 (92.2) | 1.02 (0.95–1.09) |
|  |  | Clinical improvement | 174 (77.0) | 86 (78.2) | 88 (75.9) | 0.99 (0.87–1.13) |
|  |  | Recovery | 159 (70.4) | 79 (71.8) | 80 (69.0) | 1.01 (0.86–1.19) |
| Positive SARS-CoV-2 RT-PCR^1^ | | |  |  |  |  |
|  | At day 5 | | 140/192 (72.9) | 69/95 (72.6) | 71/97 (73.2) | 0.99 (0.84–1.18) |
|  | At day 10 | | 96/166 (57.8) | 49/85 (57.7) | 47/81 (58.0) | 0.99 (0.77–1.29) |
| Data are no. (%) of all patients, unless otherwise indicated. | | | |  |  |  |
| ^1^RT-PCR could not be performed in all patients. Data are no. of positive RT-PCR/no. of RT-PCR performed (% of positive RT-PCR) | | | | | | |
