## Supplementary material for "A placebo-controlled double blind trial of hydroxychloroquine in mild-to-moderate COVID-19": Statistical Analysis Plan

Hycovid
CHU ANGERS

#### STATISTICAL ANALYSIS PLAN

| Sponsor | | | |
|---|---|---|---|
| Angers University Hospital / Centre Hospitalo-Universitaire d'Angers | | | |
| **Internal Reference** | **N° CPP** | **N° NCT (ClinicalTrials.gov)** | **ANSM** |
| HYCOVID | CPP2020-03-036 / 2020-001271-33 / 20.03.24.72431 | NCT04325893 | MEDAECNAT-2020-03-00045 |
| **Title** | | | |

### HYCOVID

### Hydroxychloroquine versus placebo in COVID-19 patients at high-risk for death or intubation: prospective, multicentre, randomized, double-blind, parallel-group, superiority trial.

| Name | Function | Signature |
|---|---|---|
| Vincent DUBEE | Study Investigator Coordinator | 0 7 JUIL. 2020 |
| Pierre-Marie ROY | Steering committee Chairman | 07 Juil 2020 |
| Elsa PAROT | Methodologist | 0 7 JUIL. 2020 |
| Bruno VIELLE | Statistician | 0 7 JUIL. 2020 |
| Jean-Marie CHRETIEN | Data-manager | 0 7 JUIL. 2020 |

### LIST OF ABBREVIATIONS

| | |
|---|---|
| CHU Angers | Centre Hospitalo-Universitaire d'Angers |
| CONSORT | Consolidated Standards of Reporting Trials |
| CRA | Clinical Research Associate |
| eCRF | Electronic Case Report Form |
| ICH | International Conference of Harmonisation |
| IDRCB | Identification de la recherche biomédicale |
| INR | International normalized Ratio |
| ITT | Intention-To-Treat |
| NCT number | ClinicalTrials.gov Identifier |
| PP | Per-Protocol |
| SAE | Serious Adverse Event |
| SAP | Statistical Analysis Plan |
| 95% CI | 95% Confidence Interval |

### VERSION HISTORY

| Version | Version Date | Updated Part | Change* | Reason | Author |
|---|---|---|---|---|---|

**Change:*
*A: Addition*
*M: Modification*
*D: Deletion*

### 1 INTRODUCTION

Severe acute respiratory syndrome coronavirus-2 (SARS-CoV-2) is a new virus that emerged from China in December 2019 and has caused an international outbreak. On March 12, 2020, World Health Organization officially declared this coronavirus disease 2019 (COVID-19) a pandemic. COVID-19 symptoms are broad-ranged, varying from pauci-symptomatic upper respiratory tract infection to severe pneumonia and ultimately fatal severe acute respiratory distress syndrome. The overall case-fatality rate ranges from 3 to 4%; risk factors for severe forms include diabetes (1), hypertension (2), obesity (3), and, above all, older age (2).
Hydroxychloroquine, a chloroquine derivative commonly used in autoimmune diseases such as systemic lupus erythematosus, have been previously shown to inhibit coronaviruses replication in vitro (4,5). Several studies have reported conflicting results regarding the efficacy and safety of hydroxychloroquine in the treatment of COVID-19, reinforcing the need of a large double-blind randomized study (6–12).

The primary aim of HYCOVID trial is to evaluate the efficacy of hydroxychloroquine versus placebo on the rate of mortality and the use of invasive ventilation among patients suffering from COVID-19 who initially do not meet severity criteria but are at high risk of aggravation. The key secondary aims are to evaluate the efficacy of hydroxychloroquine versus placebo with regard to (i) patients' clinical improvement, (ii) all-cause mortality, (iii) viral carriage duration, and (iv) incidence of venous thromboembolism events. Other secondary aims are to evaluate the efficacy of hydroxychloroquine in the subgroup of patients aged 75 years or older and to evaluate the safety of hydroxychloroquine.

The objectives of this statistical analyses plan (SAP) are:

- To clearly define all variables, endpoints/outcomes, parameters, thresholds used for the statistical analyses
- To define strategies employed regarding missing data on endpoints and possibly on adjustment variables
- To describe statistical methods performed during the analyses
- To define the populations of patients used during the analyses

### 2 TRIAL DESIGN AND OBJECTIVES

#### 2.1 Type of study

- Typology: comparison of experimental treatment to placebo
- Experimental design: prospective, multicenter, randomized, double-blind, parallel-group, superiority study
- Planned intervention: Hydroxychloroquine (400 mg bid at day 0 followed by 200 mg bid for 8 days, total dose 4 g)
- Control: placebo (2 tablets bid at day 0 followed by 1 tablet bid for 8 days, total dose 20 tablets)
- NCT (clinicaltrials.gov): NCT04325893

#### 2.2 Sample size and power consideration

Based on data from COVID-19 in China, the rate of patients requiring respiratory support and/or who dead was estimated at 20% in the study population (high-risk patients). The number of patients necessary to include per group was estimated at 615 to demonstrate, under a bilateral hypothesis, an absolute difference of 6% between the two groups (relative difference of 30%) with an alpha risk of 5% and a power of 80%. Taking into account patients lost to follow-up and those who cannot be evaluated (estimated at 5%), we planned to include 1,300 patients in total.

In late April 2020 and early May 2020, in the context of marked slowing of the epidemic in France, we observed a very low rate of inclusion, making it impossible to reach the planned number of inclusions. Thus, the study was preliminary stopped by the sponsor on date May 09, 2020.

#### 2.3 Randomization

The assignment of treatments was performed according to a 1:1 ratio by means of dynamic randomization (randomization by minimization), taking eight criteria into account:

- risk of complicated COVID-19 course with five mutually exclusive categories:
  - age ≥75 years old without oxygen requirement
  - age between 60 and 74 years old with comorbidity, without oxygen requirement
  - age < 60 years old AND oxygen requirement
  - age ≥ 75 years old AND oxygen requirement
  - age between 60 and 74 years old with comorbidity AND oxygen requirement
- diagnostic criteria of positive COVID (RT-PCR or CT scan)
- initial symptoms of COVID dating back at least seven days (yes/no)
- hospitalization (yes/no)
- concomitant treatment with azithromycin (yes/no)
- concomitant treatment with antiviral drug (yes/no)
- treatment with corticosteroids (yes/no)
- center.

#### 2.4 Study design

- Population of patients who will benefit the study results: patients suffering from COVID-19 who initially do not meet severity criteria but are at high risk of aggravation
- Number of centers: 48 centers (47 in France, 1 in the Principality of Monaco),
- Study length: inclusion = 4 months; participation = 5 months
- Visit schedule:

*Table 1: Time schedule of enrolment, interventions and assessments*

| | D0 | D1 | D3 ± 1 | D5 ± 1 | D7 ± 1 | D10 ± 1 | D14 | D28 |
|---|---|---|---|---|---|---|---|---|
| Consent/inclusion/randomization | X | Obtaining consent to continue, if required | | | | | | |
| Treatment administration | X | X | X | X | X | Until D9 | | |
| Collecting sociodemographic, clinical, biological, and therapeutic data | X | | | X | | X | X | X |
| ECG | X[1] | X | X[2] | X[2] | X[2] | X[2] | | |
| Serum potassium concentration | X[1] | | | X[2] | | | | |
| Nasopharyngeal swab sample for RT-PCR | | | | X[3] | | X[3] | | |
| Biological samples for ancillary study* | X | | | X | | X | | |
| Observation and feedback on treatment | | | | | | | X | |
| Collecting adverse events and serious adverse events | X | | X | X | X | X | X | X |
| Collecting patient status on WHO Ordinal Scale for Clinical Improvement** | | | | | | | X | X |

*for participating patients

**Consultations or telephone follow-ups if the patient is not/no longer hospitalized.

[1] before inclusion

[2] for patients with treatment that increase risk of QTc prolongation and/or interact with hydroxychloroquine and/or induce hypokalaemia

[3] For patients with positive RT-PCR SARS-CoV-2 at inclusion

#### 2.5 Objectives and endpoints

##### 2.5.1 Primary objective

- To evaluate the efficacy of hydroxychloroquine versus placebo on the rate of mortality and the use of invasive ventilation among patients suffering from COVID-19 who initially do not meet severity criteria but are at high risk of aggravation. Our hypothesis is that hydroxychloroquine administration improves prognosis of non-severe patients with COVID-19, reducing the rate of severe complications.

##### 2.5.2 Primary endpoint

- The primary outcome is the composite of death, regardless of cause, and the use of intubation and invasive ventilation within 14 days (day 14) following inclusion and the start of treatment (day 0).

##### 2.5.3 Secondary objectives

- To evaluate the efficacy of hydroxychloroquine versus placebo with regard to (i) patients' clinical improvement, (ii) all-cause mortality, (iii) viral shedding duration, and (iv) incidence of venous thromboembolism events.
- To evaluate the safety of hydroxychloroquine.
- To evaluate the efficacy of hydroxychloroquine in the subgroup of patients aged 75 years or older.

##### 2.5.4 Secondary outcomes

- Efficacy secondary outcomes include the following:
  - the rate of mortality or use of invasive ventilation in the 28 days (day 28) following inclusion and the start of treatment,
  - clinical improvement using the World Health Organization (WHO) 9-point Ordinal Scale for Clinical Improvement for COVID-19 (20) at day 14 and day 28,
  - all-cause mortality at day 14 and day 28,
  - viral shedding assessed by the rate of RT-PCR positive for SARS-CoV-2 with nasopharyngeal swab samples at day 5 and day 10 (only in the population of patients with positive RT-PCR at inclusion)
  - the rate of symptomatic venous thrombo-embolic events (VTE) at day 14 and day 28. VTE should be documented and confirmed by the adjudication committee.
- Safety secondary outcome is the rate of serious adverse events at day 28. Serious adverse events will be evaluated, and their provenance will be analyzed in line with the applicable regulations.
- Secondary outcomes in the subgroup of patients aged 75 years or older are mentioned in chapter 3.8 about subgroup analyses.

### 3 STATISTICAL METHODS

#### 3.1 General considerations

The statistical analysis will be performed by the statistician of the Biostatistics and Methodology Department, Maison de la recherche, CHU d'Angers, France

The strategy for the design and the analysis will be made in compliance with the CONSORT statement (http://www.consort-statement.org/).

Sequential analysis based on the triangular test will be performed using R software (version 3.6.3) and Stata software (version 13) will be used for other analyses.

A p-value equal or less than 0.05 will be considered as statistically significant and all statistical tests will be two-sided, unless otherwise stated

#### 3.2 Missing data and adjustment on potential factors

Missing data won't be replaced, and analysis will be made on all evaluable patients. No adjustment on potential factors will be made for the primary and secondary endpoint analyses. Only subgroup analyses are planned, as described in Chapter 3.8.

#### 3.3 Protocol deviations

All protocol deviations will be listed and summarized in a blinded manner before the database lock preliminary to the final statistical analysis. Deviations will be classified as minor or major. The process will enable the population inception for the analyses (global, randomized, intention-to-treat, safety, per-protocol) (cf. flow-chart).

The description of the protocol deviations may be found in section 6 of the SAP.

#### 3.4 Patient selection, baseline demographic and clinical characteristics, end of participation

*Patient selection*

The number of eligible patients in each center, corresponding to the number of patients diagnosed with COVID-19, will be collected. For patients not included in the trial, the reason for non-inclusion will be recorded: lack of inclusion criteria, presence of non-inclusion criteria, or refusal to participate. Demographic and clinical data will not be recorded for eligible patients. Thus, no analysis will be provided to assess the degree of potential selection bias in the study enrolment process.

*Characteristics of study population*

Initial characteristics and the end of participation (dropout, loss to follow-up, per protocol) will be summarized by treatment on the **intention-to-treat population**. Only descriptive analyses will be provided (no inferential statistics).

Results will be presented as mean ± one standard deviation if the variable follows a normal distribution and median [interquartile range] in case of other distribution. For the qualitative or categorical quantitative variables, results will be presented as numbers (proportions in %).

#### 3.5 Statistical methods for *primary* endpoint

##### 3.5.1 Analysis of primary endpoint

The primary efficacy analysis will compare the rate of death or intubation at day 14 between the placebo and hydroxychloroquine arms in the intention-to-treat population. Percentages in the two groups will be compared using Chi-squared test (Fisher's exact test if required). The null hypothesis on the primary endpoint is that there is no difference between the two arms.

Due to the current exceptional circumstances, interim analyses were performed every 50 patients to compare the two groups based solely on the main evaluation criterion. The results of the interim analyses were communicated solely to the Independent Data and Safety Monitoring Board

(IDSMB). The frequency of interim analyses was adapted depending on the number of inclusions and gathering of analyzable findings based on the main evaluation criterion. At the end of each meeting, the IDSMB determined the date for the interim analysis to be performed and for the subsequent meeting. A unilateral alpha was maintained at 2.5% during these interim analyses using the unilateral triangle test. If the alternative hypothesis limit of the triangle was exceeded, the test will have concluded to the efficacy of hydroxychloroquine. If the null hypothesis limit of the triangle was exceeded, the test will have concluded to the futility of hydroxychloroquine.

The IDSMB did not recommend to stop the study before the decision of the sponsor.

##### 3.5.2 Sensitivity analyses

The primary outcome will be analyzed on the intention-to-treat population. A sensitivity analysis will be performed in the per-protocol population. Free-event survival curves will be established between day 0 and day 28 using the Kaplan-Meier method.

No further sensitivity analysis is planned.

#### 3.6 Statistical methods for *secondary* endpoints

- The rate of the composite of use of intubation or death at day 28 will be compared in both groups using the Chi-squared test (or Fisher's exact test if required).

- Clinical evolution will be described using the World Health Organization 9-point ordinal scale for clinical improvement.

The proportion of patients with each score on the ordinal scale at day 14 and 28 will be determined and graphically presented for patients with oxygen requirement at baseline and patients without oxygen requirement.
The following events will be considered:

- Absence of deterioration (stability or decrease of at least one point in the score on the ordinal scale)
- clinical improvement (decrease of at least one point in the score on the ordinal scale)
- recovery (score of 0, 1 or 2 on the ordinal scale)

Odds-ratios for each above event at day 14 and day 28, and their 95% confidence interval will be calculated. An adjustment taking account the baseline status will be made using a Mantel-Haenszel estimation of the combined odds-ratios calculated on each stratum defined by the baseline OCSI value.

- All-cause mortality at day 14 and day 28 will be compared in both groups using the Chi-squared test (or Fisher's exact test if required); survival curves will be established between day 0 and day 28 using the Kaplan-Meier method

- The rate of RT-PCR positive for SARS-CoV-2 in nasopharyngeal samples at day 5 and day 10 will be compared in both groups using the Chi-squared test (or Fisher's exact test if required).

- The incidence of symptomatic venous thrombo-embolism occurrences at day 28 will be compared in both groups using the Chi-squared test (or Fisher's exact test if required). Free-event survival curves will be established between day 0 and day 28 using the Kaplan-Meier method.

- For the ancillary study, biological markers will be recorded as an average +/- standard deviation or median and interquartile range for non-normal distributions. Averages will be compared using the Student's t-test and medians will be compared using the Mann-Whitney test.

##### 3.7 Safety specific analyses

Safety parameters (serious adverse events (SAE) according to ICH E2A definition will be listed, summarized and tabulated by treatment group by system organ class (SOC) coded into MedDRA classification, on the **safety population** The following serious AEs related to hydroxychloroquine and mentioned in the Summary of Product Characteristics (SmPC) will be specifically assessed:

- cardiac rhythm or conduction disorders,
- seizure,
- hypoglycemia,
- vision disorder,
- vomiting,
- rash and pruritus.

Only descriptive analyses will be provided (no inferential statistics).
The results will be presented will be presented as numbers (proportions in %).

##### 3.8 Subgroup analyses

Subgroup analyses will be realized, as follows:

- The primary endpoint (the composite of death, regardless of cause, and the use of intubation and invasive ventilation at day 14) will be analyzed in the following subgroups defined by :
    - Sex
    - Age ≥75 years old
    - Oxygen requirement
    - Initial symptoms of COVID dating back less than seven days
    - Concomitant treatment with azithromycin at randomization
    - Concomitant treatment with corticosteroids at randomization
    - Treatment with corticosteroids at any time during study period (day 1 – day 14)
    - Concomitant treatment with lopinavir/ritonavir at randomization
    - Treated arterial hypertension
    - Body mass index ≥ 30 kg/m²
- The proportion of positive SARS-CoV-2 RT-PCR at day 5 and day 10 will be analyzed in the following subgroups:
    - Age ≥75 years old
    - Oxygen requirement
    - Initial symptoms of COVID dating back less than seven days
    - Concomitant treatment with azithromycin at randomization
    - Concomitant treatment with corticosteroids at randomization
    - Treatment with corticosteroids at any time during the 10 days following randomization
    - Concomitant treatment with lopinavir/ritonavir at randomization

Results will be presented in each subgroup as odds-ratio with 95% confidence interval and p-value for interaction.

- The following endpoints will be analyzed in the ≥75 years-old patient subpopulation :
    - Absence of deterioration (stability or decrease of at least one point in the score on the ordinal scale)
    - clinical improvement (decrease of at least one point in the score on the ordinal scale)
    - recovery (score of 0, 1 or 2 on the ordinal scale)
    - All-cause mortality at day 14 and day 28

### 4 POPULATION TO BE ANALYZED

#### 4.1 CONSORT 2010 Flow Diagrams

*Figure 1 Flow Diagram*

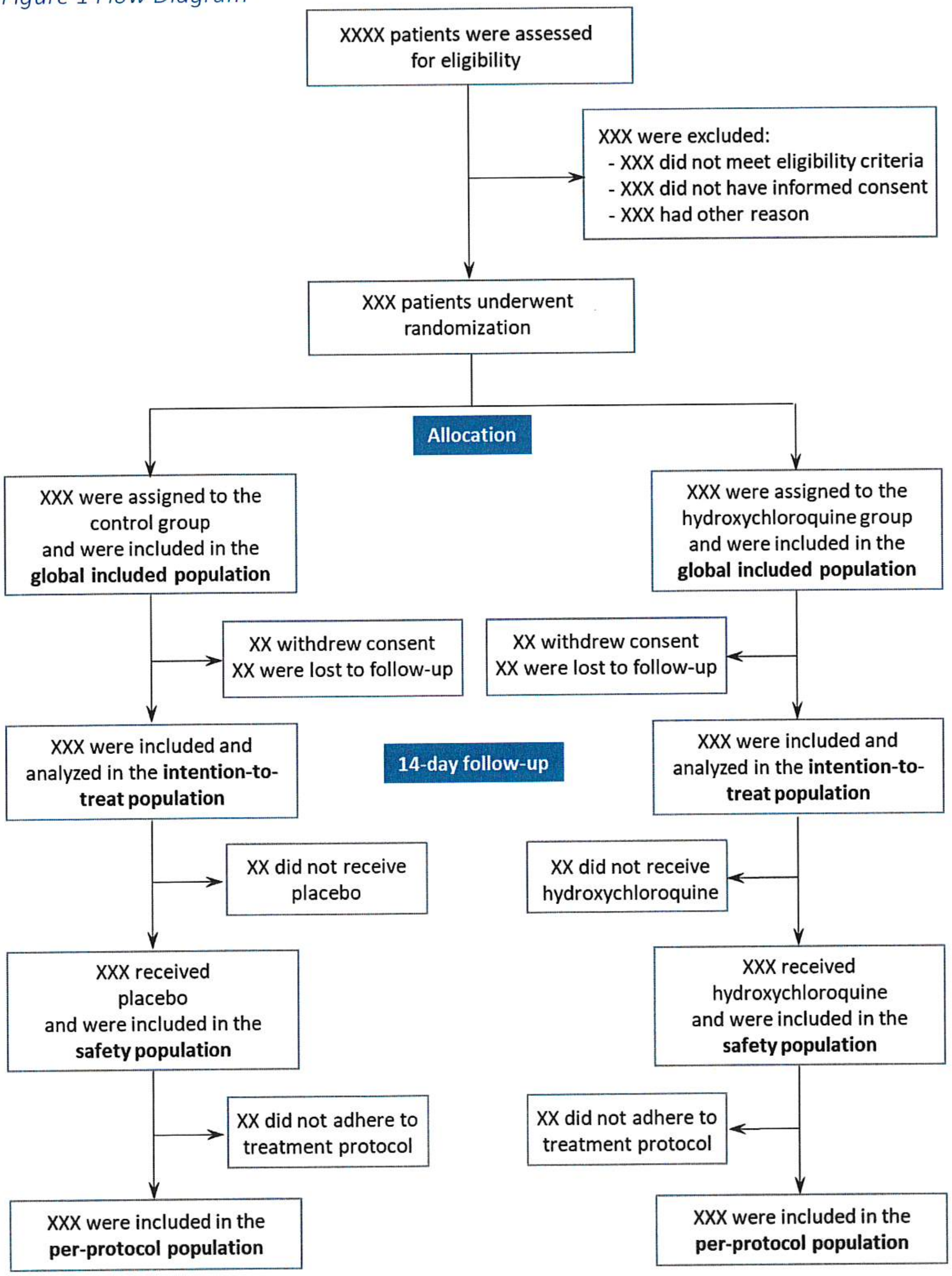

#### 4.2 Definition of populations (Table 2)

##### 4.2.1 Global included population
All included patients having a free and informed written consent adjudicated as conform.

##### 4.2.2 Intention-to-treat population
All patients having a free and informed written consent adjudicated as conform and randomized. Following the **intention-to-treat principle**, patients will be analyzed according to the treatment assigned by the randomization.

##### 4.2.3 Safety population
All randomized patients having received at least one dose of the allocated treatment

##### 4.2.4 Per-Protocol population
All randomized patients without any major protocol deviation. The major protocol deviations were pre-specified prior to unblinding treatment codes and database lock for analyses (Table 3).
They included:
- Unrespect of inclusion criteria or exclusion criteria
- Allocated treatment prematurely discontinued for another reason than clinical aggravation requiring intubation and mechanical ventilation
- Hydroxychloroquine treatment received in placebo group
- Unauthorized treatment received whatever the treatment group

#### 4.3 Populations used in analyses
The populations for statistical analysis are presented in Table 2.

*Table 1: Populations used in the analyses*

| **Population** | **Analysis** |
|---|---|
| Global set | Descriptive purpose (study inclusion flowchart) |
| Randomized set (intention-to-treat population) | Demography and clinical characteristics at baseline<br>Primary analysis of primary outcome<br>All efficacy analyses |
| Per protocol set | Sensitivity analyses |
| Safety set | Safety analyses |

### 5 DATA REVIEW

The investigator completes patient's data in an electronic Case Report Form (eCRF).
The Clinical Research Associate (CRA) visualizes the data remotely.
Data management and queries will be done under the responsibility of the CHU Angers.
A blind data review meeting will be held to validate the entire database with the investigator.
The database will be frozen for statistical analysis.
The statistical analyses will be done by the statistician of the Biostatistics and Methodology Department, Maison de la recherche, CHU Angers, France.

#### 5.1 Blind review
In order to ensure the smooth progress of the blind review meeting, some descriptions of the data will be done:
- Missing data on variables and consistencies
- Chronological order
- "Hat" variables of variables of type "if yes, please specify" or "if no, please specify"
- Missing visit

### 6 PROTOCOL DEVIATIONS

*Table 3. List of prespecified major protocol deviations.*

| Category | | Excluded from |
|---|---|---|
| A. Inclusion and exclusion criteria | | |
| A1. | Free informed consent not orally given or not signed by the patient according to local legislation | All |
| A2. | Consent for use of data withdrawn | All |
| A3. | One or more inclusion criteria lacking | PPP |
| A4. | Presence of at least one exclusion criteria* | PPP |
| B. Treatment | | |
| B1. | Allocated treatment not started | PPP and SP |
| B2. | Allocated treatment prematurely stopped for reasons other than PE met / occurrence of serious AE | PPP |
| B3. | Administration of hydroxychloroquine to a patient in the placebo group | PPP |
| B4. | Loading dose not correctly administered | PPP |
| B5. | Less than 80% of the planned doses administered, or missing data on observance with the allocated treatment | PPP |
| B6. | Improper breaking of the blind | None |
| B7. | Non-respect of the treatment regimen with a daily dose of the allocated treatment higher than expected (total number of tablets taken within less than 8 days) | PPP |
| C. Primary and secondary endpoint | | |
| C1. | Missing data on primary endpoint | ITT and PPP |
| C2. | Missing data on secondary endpoint | None** |
| D. Missing data (other than primary and secondary endpoints) | | |
| D1. | No information on concomitant treatments | None |
| D2. | No information on comorbidities | None |

PPP: leading to the exclusion from the per-protocol population analysis

ITT: leading to the exclusion from the intention-to-treat analysis

SP: leading to the exclusion from the safety population analysis

AE: adverse event

PE: primary endpoint

*Patients with negative RT-PCR on nasopharyngeal swab but with positive RT-PCR on another respiratory specimen will not be excluded from the per-protocol analysis.

**Patients with missing data on a given secondary endpoint will be excluded from the analysis of the concerned endpoint.
