## Supplementary material for "A placebo-controlled double blind trial of hydroxychloroquine in mild-to-moderate COVID-19": Members of the HYCOVID study group

**Members of the HYCOVID study group and people to be acknowledged**

### HYCOVID investigators

(last names in bold characters)

*Angers University Hospital*

Antoine **Brangier**, M.D., Service de gériatrie, CHU d’Angers, Angers, France

Philippe **Codron**, M.D., Service de neurologie, CHU d’Angers, Angers, France

Jean Michel **Lemée**, M.D., Service de neurochirurgie, CHU d’Angers, Angers, France

Virginie **Pichon**, M.D., Service de neurologie, CHU d’Angers, Angers, France

Robin **Dhersin**, Service de maladies infectieuses et tropicales, CHU d’Angers, Angers, France

*Cholet Hospital*

Roxane **Courtois**, M.D., Service de Médecine post-urgences – Maladies infectieuses, CH Cholet, Cholet, France

*Laval Hospital*

Hélène **Danielou**, M.D., Service de Médecine interne, hématologie, maladies infectieuses, CH Laval, Laval, France

Jonathan **Lebreton**, M.D., Service de Médecine interne, hématologie, maladies infectieuses, CH Laval, Laval, France

Rémi **Vatan**, M.D., Service de Médecine interne, hématologie, maladies infectieuses, CH Laval, Laval, France

*Le Mans Hospital*

Nicolas **Crochette**, M.D., Service de maladies infectieuses et tropicales, CH Le Mans, Le Mans, France

Jean-Baptiste **Lainé**, M.D., Service de maladies infectieuses et tropicales, CH Le Mans, Le Mans, France

Lucia **Perez**, M.D., Service de maladies infectieuses et tropicales, CH Le Mans, Le Mans, France

Sophie **Blanchi**, M.D., Service de maladies infectieuses et tropicales, CH Le Mans, Le Mans, France

Hikombo **Hitoto**, M.D., Service de maladies infectieuses et tropicales, CH Le Mans, Le Mans, France

*Tours University Hospital*

Louis **Bernard**, M.D., Ph.D., Service de maladies infectieuses et tropicales, CHU Tours, Tours, France

François **Maillot**, M.D., Ph.D., Service de médecine interne, CHU Tours, Tours, France

Sylvain **Marchand Adam**, M.D., Service de médecine interne, CHU Tours, Tours, France

*Quimper Hospital*

Jean-Philippe **Talarmin**, M.D., Service de médecine interne, maladies du sang et infectiologie, CH de Quimper, Quimper, France

*La Roche sur Yon Hospital*

Emeline **Gaigneux**, M.D., Service de Rhumatologie, Centre Hospitalier Départemental Vendée, La Roche sur Yon, France

Pauline **Motte-Vincent**, M.D., Service de Médecine Post-urgence, Centre Hospitalier Départemental Vendée, La Roche sur Yon, France

Marine **Morrier**, M.D., Service de Médecine Post-urgence, Centre Hospitalier Départemental Vendée, La Roche sur Yon, France

Dominique **Merrien**, M.D., Service de Médecine Post-urgence, Centre Hospitalier Départemental Vendée, La Roche sur Yon, France

Yves **Bleher**, M.D., Service de Médecine Post-urgence, Centre Hospitalier Départemental Vendée, La Roche sur Yon, France

Maxime **Flori**, M.D., Service de Cardiologie, Centre Hospitalier Départemental Vendée, La Roche sur Yon, France

Amélie **Ducet-Boiffard**, M.D., Service d’endocrinologie - diabétologie, Centre Hospitalier Départemental Vendée, La Roche sur Yon, France

Orane **Colin**, M.D., Service de Médecine Post-urgence, Centre Hospitalier Départemental Vendée, La Roche sur Yon, France

Ronan **Février**, M.D., Service de Médecine Gériatrique, Centre Hospitalier Départemental Vendée, La Roche sur Yon, France

*Tourcoing Hospital*

Pauline **Thill**, MD, Service Universitaire des Maladies Infectieuses et du Voyageur, CH de Tourcoing, Tourcoing, France

Macha **Tetart**, MD, Service Universitaire des Maladies Infectieuses et du Voyageur, CH de Tourcoing, Tourcoing, France

François **Demaeght**, MD, Service Universitaire des Maladies Infectieuses et du Voyageur, CH de Tourcoing, Tourcoing, France

Barthelemy **Lafond-Desmurs**, MD, Service Universitaire des Maladies Infectieuses et du Voyageur, CH de Tourcoing, Tourcoing, France

Maxime **Pradier**, MD, Service Universitaire des Maladies Infectieuses et du Voyageur, CH de Tourcoing, Tourcoing, France

Agnes **Meybeck**, MD, Service Universitaire des Maladies Infectieuses et du Voyageur, CH de Tourcoing, Tourcoing, France

Marjorie **Picaud**, MD, Service de pneumologie, CH de Tourcoing, Tourcoing, France

*Orléans Hospital*

Thierry **Prazuck**, M.D., Service de maladies infectieuses et tropicales, CH régional d’Orléans, Orléans, France

*Nantes University Hospital*

Guillaume **Chapelet**, M.D., Service de médecine aiguë gériatrique, CHU de Nantes, Nantes, France

Agnès **Rouaud**, M.D., Service de médecine aiguë gériatrique, CHU de Nantes, Nantes, France

Paul **Le Turnier**, M.D., Service de maladies infectieuses et tropicales, CHU de Nantes, Nantes, France

*Niort Hospital*

Simon **Sunder**, M.D., Service de maladies infectieuses et tropicales, CH de Niort, Niort, France

*Lorient Hospital*

Aurélien **Lorleac'h**, M.D., Service de médecine interne - maladies infectieuses, Hôpital du Scorff, Groupe Hospitalier Bretagne Sud, Lorient, France

Christophe **Dollon**, M.D., Service de médecine polyvalente, Hôpital du Scorff, Groupe Hospitalier Bretagne Sud, Lorient, France

Antoine **Jacquet**, M.D., Service de médecine polyvalente, Hôpital du Scorff, Groupe Hospitalier Bretagne Sud, Lorient, France

Francois **Le Vely**, M.D., Service de médecine polyvalente, Hôpital du Scorff, Groupe Hospitalier Bretagne Sud, Lorient, France

*Brest University Hospital*

Pierre **Gazeau**, M.D., Service de maladies infectieuses et tropicales, CHU de Brest, Brest, France

Séverine **Ansart**, M.D., Ph.D., Service de maladies infectieuses et tropicales, CHU de Brest, Brest, France

*Cherbourg Hospital*

Hélène **Roger**, M.D., Service de médecine interne et maladies infectieuses, CH du Cotentin, Cherbourg, France

François **Laterza**, M.D., Service de médecine interne et maladies infectieuses, CH du Cotentin, Cherbourg, France

*Saint-Brieuc Hospital*

Rodolphe **Buzelé**, M.D., Service de médecine interne et maladies infectieuses, CH Yves Le Foll, Saint-Brieuc, France

*Créteil – APHP University Hospital*

Fella **Tahmi**, M.D., Service de Gériatrie, CHU Henri Mondor – APHP, Créteil, France.

Raphael **Lepeule**, M.D., Unité Transversale de Traitement des Infections, CHU Henri Mondor – APHP, Créteil, France.

*Saint-Antoine – APHP University Hospital*

Karine **Lacombe**, M.D., Ph.D., Service de maladies infectieuses et tropicales, CHU Saint Antoine – APHP, Paris, France

Bénédicte **Lefebvre**, M.D., Service de maladies infectieuses et tropicales, CHU Saint Antoine – APHP, Paris, France

*Saint-Etienne University Hospital*

Thomas **Célarier**, M.D., Service de gérontologie clinique, CHU Saint-Etienne, Saint-Etienne, France

Amandine **Gagneux-Brunon**, M.D., PhD., Service d’Infectiologie, CHU Saint-Etienne, Saint-Etienne, France

Elisabeth **Botelho-Nevers**, M.D., Ph.D., Service d’infectiologie, CHU Saint-Etienne, Saint-Etienne, France

*Toulouse University Hospital*

Marc **Bernard**, M.D., Post-Urgences Médicale, CHU Toulouse, Toulouse, France

Camille **Garnier**, M.D., Service de maladies infectieuses et tropicales, CHU Toulouse, Toulouse, France

Morgane **Mourguet**, M.D., Service de maladies infectieuses et tropicales, CHU Toulouse, Toulouse, France

Gregory **Pugnet**, M.D., Ph.D, Service de Médecine interne, CHU Toulouse, Toulouse, France

Sara **Vienne-Noyes**, M.D., Post-Urgences Gériatrique, CHU Toulouse, Toulouse, France

Guillaume **Martin-Blondel**, M.D., Ph.D., Service de maladies infectieuses et tropicales, CHU Toulouse, Toulouse, France

Pierre **Delobel**, M.D., Ph.D., Service de maladies infectieuses et tropicales, CHU Toulouse, Toulouse, France

Gaspard **Grouteau**, M.D., Service de maladies infectieuses et tropicales, CHU Toulouse, Toulouse, France

Alexa **Debard**, M.D., Service de maladies infectieuses et tropicales, CHU Toulouse, Toulouse, France

Laurent **Guilleminault**, M.D., Ph.D., Service de Pneumologie, CHU Toulouse, Toulouse, France

*Melun Hospital*

Pauline **Arias**, M.D., Service de médecine polyvalente et maladies infectieuses, Groupe Hospitalier Sud Ile de France, Melun, France

Catherine **Chakvetadze**, Service de médecine polyvalente et maladies infectieuses, Groupe Hospitalier Sud Ile de France, Melun, France

Clara **Flateau**, Service de médecine polyvalente et maladies infectieuses, Groupe Hospitalier Sud Ile de France, Melun, France

Aude **Kopp**, Service de médecine polyvalente et maladies infectieuses, Groupe Hospitalier Sud Ile de France, Melun, France

*Dijon University Hospital*

Alain **Putot**, ervice de médecine interne gériatrie, CHU Dijon Bourgogne, Dijon, France

Jeremy **Barben**, Service de médecine interne gériatrie, CHU Dijon Bourgogne, Dijon, France

Suzanne **Mouries Martin**, Service de médecine interne et maladies systémiques, CHU Dijon Bourgogne, Dijon, France

Valentine **Nuss**, Service de médecine interne gériatrie, CHU Dijon Bourgogne, Dijon, France

Lionel **Piroth**, M.D., Ph.D., Département d’infectiologie, CHU Dijon Bourgogne, Dijon, France

*Princesse Grace – Monaco Hospital*

Yann-Erick **Claessens**, M.D., Ph.D., Service des urgences, Centre Hospitalier Princesse Grace, Monaco, Principauté de Monaco

*Versailles Hospital*

Veronique **Hentgen**, M.D., Service de pédiatrie, CH Versailles - Hôpital André Mignot, Le Chesnay, France

*Colmar Hospital*

Martin **Martinot**, M.D., Service de Maladies Infectieuses et Tropicales, Hôpital Pasteur Colmar, Colmar, France

Maxime **Bach-Bunner**, M.D., Service de médecine interne, Hôpital Pasteur Colmar, Colmar, France

Thomas **Bonijoly**, M.D., Service de Maladies Infectieuses et Tropicales, Hôpital Pasteur Colmar, Colmar, France

Simon **Gravier**, M.D., Service de Maladies Infectieuses et Tropicales, Hôpital Pasteur Colmar, Colmar, France

Jean-Marc **Michel**, M.D., Service de gériatrie, Hôpital Pasteur Colmar, Colmar, France

*Agen-Nerac Hospital*

Mathilde **Andreu**, M.D., Service de pneumologie, CH Agen, Agen, France

Mélanie **Roriz**, M.D., Service de médecine interne, CH Agen, Agen, France

*Caen University Hospital*

Aurélie **Baldolli**, M.D., Service de Maladies Infectieuses et Tropicales, CHU Caen, Caen, France

*Saint-Nazaire Hospital*

Julia **Brochard**, M.D., Service de médecine polyvalente – infectiologie, CH Saint Nazaire, St Nazaire, France

*Nantes – Confluent Hospital*

Olivier **Grossi**, M.D., Service de médecine interne et maladies infectieuses, Hôpital privé du Confluent, Nantes, France

Samuel **Pineau**, M.D., Service de médecine interne et maladies infectieuses, Hôpital privé du Confluent, Nantes, France

*Limoges University Hospital*

Josselin **Brisset**, M.D., Service de maladies infectieuses et tropicales, CHU de Limoges, Limoges, France

Edouard **Desvaux**, M.D., Service de médecine interne et gériatrique, CHU de Limoges, Limoges, France

Guillaume **Gondran**, M.D., Service de médecine interne A, CHU de Limoges, Limoges, France

Jean-François **Faucher**, M.D., Ph.D., Service de maladies infectieuses et tropicales, CHU de Limoges, Limoges, France

Paul-Antoine **Quesnel**, M.D., Service d’accompagnement et soins palliatifs, CHU de Limoges, France

Holy **Bezanahary**, M.D., Service de médecine interne A, CHU de Limoges, Limoges, France

Clément **Danthu**, M.D., Service de néphrologie, CHU de Limoges, Limoges, France

Blandine **Gutierrez**, M.D., Service de médecine interne A, CHU de Limoges, Limoges, France

Kim **Ly**, M.D.,Ph.D., Service de médecine interne A, CHU de Limoges, Limoges, France

Yannick **Simonneau**, Service de pathologie respiratoire, CHU de Limoges, Limoges, France

Anne **Cypierre**, M.D., Service de maladies infectieuses et tropicales, CHU de Limoges, Limoges, France

Pauline **Pinet**, M.D., Service de maladies infectieuses et tropicales, CHU de Limoges, Limoges, France

Hélène **Durox**, M.D., Service de maladies infectieuses et tropicales, CHU de Limoges, Limoges, France

Sophie **Ducroix-Roubertou**, M.D., Service de maladies infectieuses et tropicales, CHU de Limoges, Limoges, France

Claire **Genet**, M.D., Service de maladies infectieuses et tropicales, CHU de Limoges, Limoges, France

*Poitiers University Hospital*

Guillaume **Beraud**, M.D., Service de maladies infectieuses et tropicales, CHU de Poitiers, Poitiers, France

Gwenael **Le Moal**, M.D., Service de maladies infectieuses et tropicales, CHU de Poitiers, Poitiers, France

Blandine **Rammaert**, M.D., Ph.D., Service de maladies infectieuses et tropicales, CHU de Poitiers, Poitiers, France

*Amiens University Hospital*

Jean-Philippe **Lanoix**, M.D., Ph.D., Service de Service de maladies infectieuses et tropicales, CHU d’Amiens, Amiens, France

Claire **Andrejak**, M.D., Ph.D., Service de pneumologie, CHU Amiens, Amiens, France

Cédric **Joseph**, M.D., Service de Service de maladies infectieuses et tropicales, CHU d’Amiens, Amiens, France

Sandrine **Soriot-Thomas**, M.D., Centre de recherche clinique, CHU d’Amiens, Amiens, France

*Bobigny – APHP University Hospital*

Robin **Dhote**, M.D., Ph.D., Service de médecine interne, CHU Avicennes – APHP, Bobigny, France

Sébastien **Abad**, M.D., Service de médecine interne, CHU Avicennes – APHP, Bobigny, France

Ruben **Benainous**, M.D., Service de médecine interne, CHU Avicennes – APHP, Bobigny, France

*Cergy-Pontoise Hospital*

Jean-François **Boitiaux**, M.D., Service de pneumologie, CH René-Dubos, Cergy-Pontoise, France

Guillaume **Briend**, M.D., Service de pneumologie, CH René-Dubos, Cergy-Pontoise, France

Celine **Gonfroy**, M.D., Service d’endocrinologie, CH René-Dubos, Cergy-Pontoise, France

Stanislas **Harent**, M.D., Service de médecine interne et maladies infectieuses, CH René-Dubos, Cergy-Pontoise, France

Aurore **Lagrange**, M.D., M.D., Service de pneumologie, CH René-Dubos, Cergy-Pontoise, France

*Valencienne Hospital*

Alina **Tone**, M.D., Service d’infectiologie, CH de Valencienne, Valencienne, France

*Valencienne – Clinique Tessier Hospital*

Thomas **Gey**, M.D., M.D., Service de pneumologie, Clinique Teissier, Valencienne, France

*Henri-Mondor – APHP University Hospital*

Andrea **Toma**, M.D., Ph.D., Service A1, Hôpital Dupuytren, CHU Henri Mondor-APHP, Draveil, France

Amaury **Broussier**, M.D., Service de Gériatrie Ambulatoire, Hôpital Emile Roux – GHU-APHP Henri Mondor, Limeil Brévannes, France

Sandrine **Etienne**, M.D., Service de Gérontologie 1, Hôpital Emile Roux – GHU-APHP Henri Mondor, Limeil Brévannes, France

Yann **Spivac**, M.D., Service de Gérontologie 1, Hôpital Emile Roux – GHU-APHP Henri Mondor, Limeil Brévannes, France

*Chalon-sur-Saône Hospital*

Benoit **Martha**, M.D., Service de maladies infectieuses, CH de Chalon-sur-Saône, Chalon-sur-Saône, France

Nathalie **Roch**, M.D., Service de maladies infectieuses, CH de Chalon-sur-Saône, Chalon-sur-Saône, France

Pierre **Diaz**, M.D., Service de pneumologie, CH de Chalon-sur-Saône, Chalon-sur-Saône, France

Danièle **N’guyen Baranoff**, M.D., Service de pneumologie, CH de Chalon-sur-Saône, Chalon-sur-Saône, France

*Marseille European Hospital*

Stanislas **Rebaudet**, M.D., Ph.D., Hôpital Européen, Marseille, France

*Auxerre Hospital*

François **Jourda**, M.D., Service de Cardiologie, CH d’Auxerre, Auxerre, France

*Diaconnesses Croix-Saint-Simon Hospital*

Valérie **Zeller**, M.D., Service de Médecine interne et infectiologie, Groupe Hospitalier Diaconesses – Croix Saint Simon, Paris, France

*Marseille – Saint Joseph Hospital*

Boris **Bienvenu**, M.D., Ph.D., Service de médecine interne, Hôpital Saint Joseph, Marseille, France

Arnaud **Boyer**, M.D., Service de pneumologie, Hôpital Saint Joseph, Marseille, France

### Composition of the HYCOVID management team

***Steering committee (authors)***

Isabelle Pellier

Alain Mercat

Astrid Darsonval

Odile Blanchet

Marc-Antoine Custaud

Caroline Lefeuvre

Elsa Parot-Schinkel

Bruno Vielle

Marie Briet

Pierre-Marie Roy (Chair)

Vincent Dubée

***Independant data safety and monitoring board***

Bertrand Guidet (Chair)

Patrick Mismetti

Eric Vicaut

***Independent adjudication of clinical events committee***

Olivier Sanchez

Philippe Girard

Antoine Elias

Francis Couturaud

***Study management***

*Coordination*

Béatrice Gable

Sybille Lazareff

Loïc Carballido

Catherine Hue

*Data management*

Jean-Marie Chrétien

Adrien Goraguer

Lucie van Eeckhoutte

### Acknowledgments

Other people who contributed to the study but do not fulfill the criteria for auhtorship

| **Center** | **First Name** | **Last Name** | **Role/grade** |
| --- | --- | --- | --- |
| Agen | Amina | El Arabi | Research management assistant |
| Agen | Albert | Trinh-Duc | Doctor of Medicine |
| Angers | Adele | Nseka | Clinical Research Assistant |
| Angers | Adrien | Goraguer | Data manager |
| Angers | Amélie | Ifrah | Doctor of Pharmacy |
| Angers | Amine | Amroun | Clinical Research Assistant |
| Angers | Anne-Lise | Gravouille | Clinical Research Assistant |
| Angers | Anne-Lyse | Quibel | Clinical Research Assistant |
| Angers | Aurélie | Jamet | Doctor of Pharmacy |
| Angers | Béatrice | Gable | Clinical Study Coordinator |
| Angers | Catherine | Hue | Doctor of Pharmacy |
| Angers | Cécile | Hervé | Data manager |
| Angers | Cédric | Annweiler | Doctor of Medicine |
| Angers | Celina | Pruvost | Clinical Research Assistant |
| Angers | Chloe | Ragueneau | Clinical Research Assistant |
| Angers | Christophe | Laplace | Clinical Research Assistant |
| Angers | Dominique | Boosz | Clinical Research Assistant |
| Angers | Elise | Houssin | Data manager |
| Angers | Emmanuel | Quemener | Clinical Research Assistant |
| Angers | Estelle | Perrin | Clinical Research Assistant |
| Angers | Henri | Guepin | Clinical Research Assistant |
| Angers | Ines | Aoudia | Clinical Research Assistant |
| Angers | Jean-Marie | Chrétien | Data manager |
| Angers | Julien | Dolou | Clinical Research Assistant |
| Angers | Justine | Gonsard | Clinical Study Coordinator |
| Angers | Loïc | Carballido | Research team director |
| Angers | Lucie | Van Eeckhoutte | Data manager |
| Angers | Marie | Barbe | Clinical Study Coordinator |
| Angers | Marine | Thomas | Clinical Research Assistant |
| Angers | Patricia | Gougeon | Clinical Research Assistant |
| Angers | Pierre | Abgueguen | Doctor of Medicine |
| Angers | Pierre-Marie | Le Meur | Data manager |
| Angers | Sami | Rehaiem | Clinical Research Assistant |
| Angers | Samuel | Thiery | Clinical Research Assistant |
| Angers | Sebastien | Bivaud | Clinical Research Assistant |
| Angers | Sébastien | Bivaud | Clinical Research Assistant |
| Angers | Sophie | Thayanithy | Clinical Research Assistant |
| Angers | Stephanie | Legrand | Clinical Research Assistant |
| Angers | Sybille | Lazareff | Clinical Study Coordinator |
| Angers | Valérie | Ugo | Doctor of Medicine |
| Angers | Wojciech | Trzepizur | Doctor of Medicine |
| Caen | Laure | Allain | Clinical Research Assistant |
| Caen | Severine | Gautier | Clinical Research Assistant |
| Caen | Sylvie | Brucato | Clinical Research Assistant |
| Chalon/Saône | Claudine | Desbrosses | Research management assistant |
| Chalon/Saône | Jérôme | Poirot | Research management assistant |
| Chalon/Saône | Jérôme | Coutet | Doctor of Pharmacy |
| Cherbourg | Aude | Grébert-Manuardi | Clinical Study Coordinator |
| Cherbourg | Bertrand | Sauneuf | Doctor of Medicine |
| Cherbourg | Bleuenn | Dreves | Doctor of Medicine |
| Cherbourg | Caroline | Viron | Doctor of Medicine |
| Cherbourg | Cécilia | Deac | Doctor of Medicine |
| Cherbourg | Cindy | Beche | Doctor of Medicine |
| Cherbourg | Eleonore | Deck | Doctor of Medicine |
| Cherbourg | Thierry | Colin | Doctor of Medicine |
| Cholet | Caroline | Francois | Research management assistant |
| Cholet | Laetitia | Seguin | Research management assistant |
| Cholet | Laura | Vallee | Research management assistant |
| Cholet | Marie | Gaume | Doctor of Pharmacy |
| Colmar | Anne | Pachart | Research management assistant |
| Colmar | Sabine | Camara | Research management assistant |
| Dupuytren | Patrick | Leglise | Doctor of Pharmacy |
| Dupuytren | Zafy | Ralantonisainana | Doctor of Medicine |
| Emile Roux | Veronique | Vianefe | Research management assistant |
| GH Croix Saint Simon | Youbes | Kerroumi | Clinical Research Assistant |
| Hôpital Européen Marseille | Amélie | Rognon | Doctor of Pharmacy |
| Hôpital Européen Marseille | Christina | Psomas | Doctor of Medicine |
| Hôpital Européen Marseille | Corine | Madjarian | Clinical Research Assistant |
| Hopital Privé du Confluent | Claire | Hussenet | Doctor of Medicine |
| Hopital Privé du Confluent | Delphine | Egret | Research management assistant |
| La Roche sur Yon | Hélène | Pélerin | Research management assistant |
| La Roche sur Yon | Catherine | Albrecht | Research management assistant |
| La Roche sur Yon | Edwige | Migne | Clinical Research Nurse |
| La Roche sur Yon | Hélène | Durand | Clinical Research Nurse |
| La Roche sur Yon | Peguy | Chupeau | Clinical Research Nurse |
| Le Chesnay | Beulaygue | Anais | Research management assistant |
| Le Chesnay | Greder Belan | Alix | Doctor of Medicine |
| Le Chesnay | Kaoudji | Salima | Research management assistant |
| Limoges | Eloïse | Dobbels | Research management assistant |
| Limoges | Françoise | Renon-Carron | Doctor of Pharmacy |
| Limoges | Hugues | Caly | Doctor of Medicine |
| Limoges | Laurent | Fourcade | Doctor of Medicine |
| Limoges | Lynda | Pervieux | Research management assistant |
| Lorient | Mariella | Le Saux | Research management assistant |
| Melun | Tracy | Youbong | Doctor of Medicine |
| Mondor | Amel | Gouja | Research management assistant |
| nantes | Albane | Soria | Clinical Research Assistant |
| Nantes | Benjamin | Gaborit | Doctor of Medicine |
| Nantes | Ernesto | Paredes | Clinical Research Assistant |
| Nantes | Jérémie | Orain | Clinical Research Assistant |
| Nantes | Morane | Cavellec | Clinical Research Assistant |
| Nantes | Morgane | Le Bras | Clinical Research Assistant |
| Niort | Anabele | Dos Santos | Doctor of Medicine |
| Niort | Diane | Chuillet Moreau | Clinical Research Assistant |
| Niort | Jeanne | Oddoz | Clinical Research Assistant |
| Niort | Kinda | Schepers | Doctor of Medicine |
| Niort | Veronique | Goudet | Doctor of Medicine |
| Pontoise | Celeste | Lambert | Doctor of Medicine |
| Pontoise | Marine | Gosset | Doctor of Medicine |
| Pontoise | Sixtine | Decaux | Resident |
| Quimper | Clémence | Arrivé | Clinical Research Assistant |
| Quimper | Lydie | Khatchatourian | Doctor of Medicine |
| Quimper | Marie-Sarah | Fangous | Doctor of Pharmacy |
| Quimper | Nadia | Saïdani | Doctor of Medicine |
| Quimper | Nicolas | Cassou | Doctor of Pharmacy |
| Quimper | Pascaline | Rameau | Clinical Research Assistant |
| Saint Brieuc | Marie Cécile | Hervé | Research management assistant |
| Saint-Antoine Paris | Christian | Tran | Clinical Research Assistant |
| Saint-Antoine Paris | Cyrielle | Le Taillandier | Clinical Research Assistant |
| Saint-Antoine Paris | Jean-Luc | Lagneau | Clinical Research Assistant |
| Saint-Antoine Paris | Julie | Lamarque | Clinical Research Assistant |
| Saint-Antoine Paris | Juliette | Blondy | Clinical Research Assistant |
| Saint-Antoine Paris | Manuela | Le Cam | Clinical Research Assistant |
| Saint-Etienne | Anne | Pouvaret | Doctor of Medicine |
| Saint-Etienne | Anne | Fresard | Doctor of Medicine |
| Saint-Etienne | Céline | Cazorla | Doctor of Medicine |
| Saint-Etienne | Cyrille | Renaud | Clinical Research Assistant |
| Saint-Etienne | Maelle | Detoc | Clinical Research Assistant |
| Saint-Etienne | Marie France | Lutz | Doctor of Medicine |
| Saint-Etienne | Veronique | Ronat | Clinical Research Assistant |
| St Nazaire | Aurore | Deininger | Doctor of Pharmacy |
| St Nazaire | Pauline | Le Floch | Clinical Research Assistant |
| St Nazaire | Servane | Vastral | Clinical Research Assistant |
| Tourcoing | Marie-Caroline | Marien | Clinical Research Assistant |
| Tourcoing | Pauline | Cornavin | Clinical Research Assistant |
| Tourcoing | Solange | Trehoux | Clinical Research Assistant |
| Tourcoing | Sylvie | Devlieger | Clinical Research Assistant |
| Tourcoing | Vincent | Derdour | Clinical Research Assistant |
